# Mosaic Loss of Y chromosome associates with lung function, emphysema and epigenetic aging

**DOI:** 10.1101/2025.07.30.25332379

**Authors:** Woei-Yuh Saw, Kangjin Kim, Yichen Huang, Jeong H. Yun, Xiaolong Ma, Jason Bacon, Yash Pershad, Daniel Levy, George T. O’Connor, Eric Boerwinkle, R. Graham Barr, Stephen S. Rich, Jerome I. Rotter, April P. Carson, Laura M. Raffield, Sina A. Gharib, Traci M. Bartz, Bruce M. Psaty, Tamar Sofer, Kari E. North, Robert Kaplan, Elizabeth C. Oelsner, Ani Manichaikul, Alexander G. Bick, Paul Sheet, Alexander P. Reiner, NHLBI Trans-Omics for Precision Medicine Consortium, Yasminka A. Jakubek, Paul L. Auer, Michael H. Cho, Dawn L. DeMeo

## Abstract

Mosaic loss of Y chromosome (mLOY) in blood cells is an age-related somatic mutation, but its relationship with pulmonary health remains undercharacterized. Leveraging mLOY assessment in over 12,000 men, including 5,097 from the COPDGene Study and 7,235 from six additional cohorts in Trans-Omics for Precision Medicine program, we investigated its association with respiratory outcomes and epigenetic aging. Cross-sectionally, mLOY was associated with airflow obstruction with prevalence increasing with age, particularly in men with a former smoking history. Longitudinally, mLOY associated with lung function decline. Notably, mLOY was also associated with greater CT-quantified lung emphysema and faster pace epigenetic aging. Prospectively, in participants with normal lung function at baseline, mLOY was associated with lower future lung function and faster pace of epigenetic aging. These associations remained robust after adjusting for clonal hematopoiesis and telomere length. Collectively, these findings position mLOY as a potential biomarker of respiratory aging and obstructive lung disease.

**Graphical Abstract:** 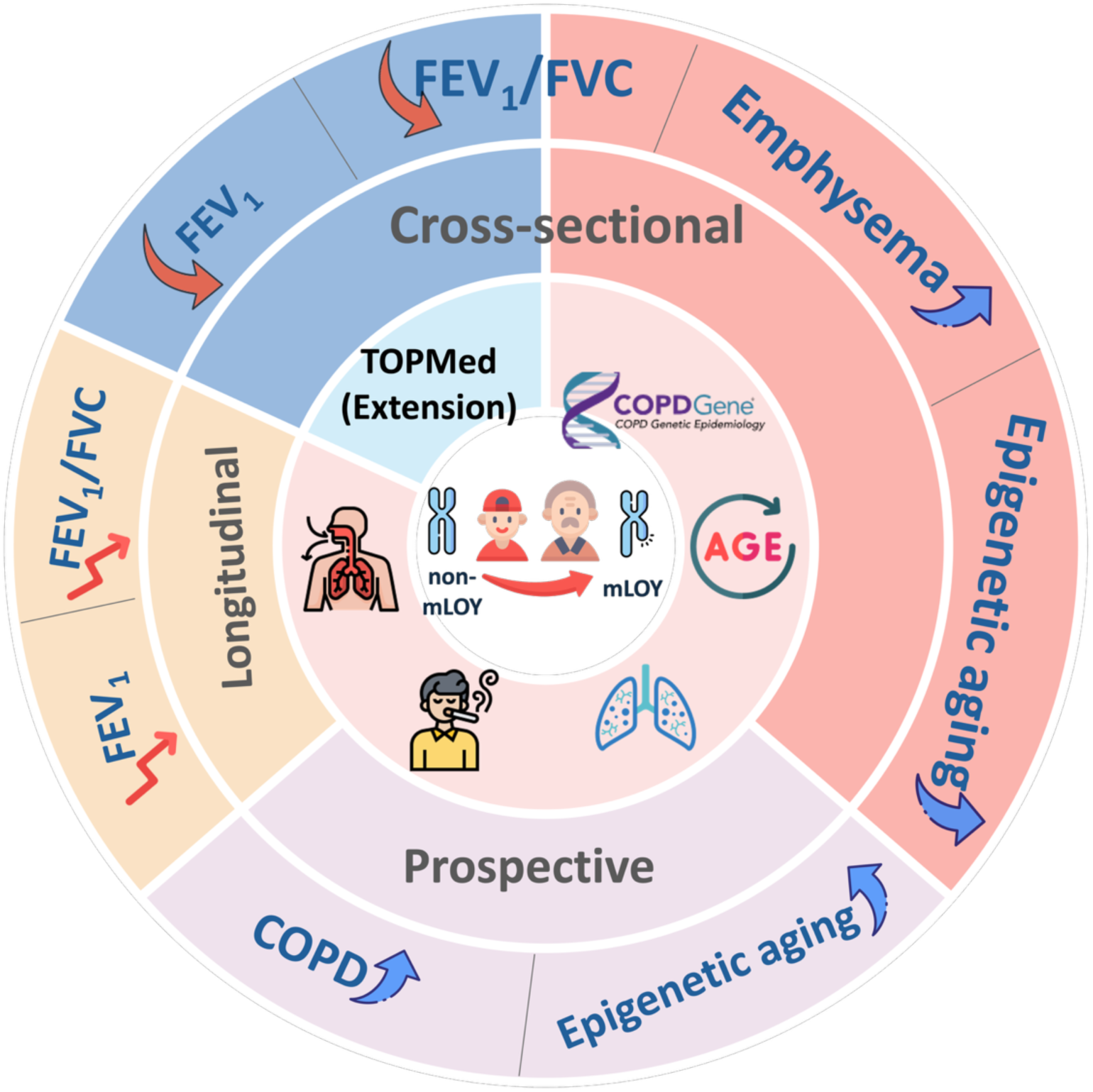

## Introduction

Aging is a major risk factor for many lung diseases, and accelerated lung aging may drive lung disease progression^1^. The Y chromosome has gained increasing attention for its role in human health beyond primary sex determination. Emerging evidence suggests that the mosaic loss of Y chromosome (mLOY) in somatic cells, a phenomenon observed in men as they age, may be associated with adverse health outcomes, including increased mortality and a higher risk of age-related non-communicable diseases such as Alzheimer disease, cardiovascular disease, type II diabetes, chronic kidney disease, and atrial fibrillation^2–6^. This mosaicism has also been associated with cigarette smoking, which is a significant factor contributing to both lung disease and mLOY, in addition to age^7,8^.

Chronic Obstructive Pulmonary Disease(COPD) represents a major global health burden increasing with aging of the population^9^. Smoking is also a well-established cause of lung function impairment and a major contributor to COPD^7^. Men with a history of smoking may therefore face a dual burden, through the direct effects of smoking on lung health and a higher prevalence of mLOY as they age. The epidemiological relationships between mLOY, lung function, emphysema, and COPD require further clarification.

Data from the UK Biobank (UKB) suggest that mLOY is associated with an increased risk of COPD, even after adjusting for varying levels of smoking intensity^10^. Additionally, recent findings from the China Kadoorie Biobank(CKB) reveal that mLOY is associated with increased risks of lung cancer, COPD, and idiopathic pulmonary fibrosis (IPF), with the highest risks observed among current smokers with mLOY^11^. Although positive interactions between mLOY and smoking were observed in the UKB, similar risk patterns in the CKB cohort suggest that having both mLOY and current smoking may amplify disease risk more than either factor alone^11^. However, spirometric measures of lung function and quantitative CT measures of emphysema were not included in these analyses. Experimental evidence also supports mLOY’s potential role in lung disease. Accelerated fibrotic responses have been observed in the lung interstitium of mice with mLOY^6^, suggesting a possible contribution to pulmonary fibrosis. However, some evidence suggests that mLOY may not be a major driver of IPF^12^. Overall, further in-depth research is needed regarding the relation of mLOY to lung disease in men.

Here, we investigate the association between mLOY and cross sectional, longitudinal and prospective respiratory outcomes, including spirometry, quantitative emphysema and COPD. By focusing on population-level data, this study aims to explore the relationship between mLOY and lung health, providing a new perspective on this somatic mutation that may portend respiratory outcomes in aging men.

## Methods and Materials

### Discovery study population

This study leverages the COPDGene^13^ cohort, a 21-center prospective cohort designed to investigate genetic factors related to susceptibility to and severity of COPD. Participant visits occurred approximately every 5 years (Phase 1 baseline enrolment (2007-2011), Phase 2 (2013-2017) and Phase 3 (2018-2023)). Detailed descriptions of the cohort, recruitment methods, and data collection procedures have been published^13^. Exclusion criteria were pre-existing interstitial lung disease, bronchiectasis and other chronic lung disease, except for asthma. Blood samples were collected for DNA extraction and individual baseline for phenotype data was defined with reference to the date of the blood collection used for whole genome sequencing (WGS) (Phase 1 or Phase 2). For this analysis, we included 5,097 male participants with sex verified with WGS. Comprehensive questionnaires detailing respiratory symptoms, medical history, demographic information, spirometry and Computed Tomography (CT) exam of the chest were collected at each study visit. Spirometry was performed according to ATS standards^14^. All participants provided written informed consent, and each study center obtained institutional review board approval.

### Lung function and emphysema outcomes

Lung function traits analysed included the volume of air exhaled in the first second of a forced breath (Forced Expiratory Volume in one second, FEV_1_), the total volume exhaled during a forced breath (Forced Vital Capacity, FVC), and the ratio of these two measures (FEV_1_/FVC). Lung function was assessed both before and after administration of a bronchodilator medication.

COPD was classified according to the Global Initiative for Chronic Obstructive Lung Disease (GOLD) criteria^15^. All cases of COPD were defined by FEV_1_/FVC ≤ 0.7. Three groups were evaluated: all grades of COPD, from mild to very severe (grades 1 to 4); moderate to severe COPD (grades 2 to 4); and severe disease only (grades 3 and 4). Controls had normal spirometry (FEV_1_/FVC >0.7 and FEV_1_ ≥ 80 percent of the predicted value). Individuals with Preserved Ratio Impaired Spirometry (PRSIm), normal FEV_1_/FVC but a reduced FEV_1_, were analysed as a separate group.

Percent predicted lung function values were calculated using reference equations from the Global Lung Function Initiative (GLI)^16^. Inspiratory and expiratory chest computed tomography scans were obtained following a standardized imaging protocol. Scanner type was included as a covariate in analyses of quantitative imaging measures. Emphysema (LAA950) was assessed using the percentage of low-attenuation areas ≤ −950 Hounsfield Units (HU) on inspiratory computed tomography scans. Quantitative analysis of imaging was performed by Thirona (Nijmegen, Netherlands).

### Pace of Aging

DNA methylation data was generated for COPDGene using the Illumina EPIC V1 array according to standard laboratory protocols. DNA methylation data were available from samples matched to the WGS timepoint. Epigenetic pace of aging was estimated using the DunedinPoAm38^17^ methylation score, where a higher score suggests a faster pace of aging. The DunedinPoAm38 score was calculated using the ‘methylCIPHER’^18^ R package.

### Whole genome sequencing and mLOY classification

WGS data utilized in this study were generated through the NIH NHLBI Trans-Omics for Precision Medicine(TOPMed) program^19^. WGS of DNA was performed at an average depth of 38X using Illumina X10 technology at two sequencing centers (Broad Genomics and Washington University). Details of the quality control procedures have been published^19^. Genetic ancestry principal components (PCs) were generated from the WGS data.

Mosaic loss of Y chromosome (mLOY) detection was performed using allele-specific read depth data from WGS. The Mosaic Chromosomal Alterations^20,21^ (MoChA v1.15) software was employed to identify allelic imbalances and reduced coverage in pseudoautosomal region 1 (PAR1), which are suggestive of mLOY. MoChA analysis included filtering options to improve the accuracy of the mLOY calls. Cell fraction (CF) estimates for mLOY were calculated using the MoChA algorithm, as described in Jakubek et al^22,23^; mLOY status was dichotomized based on CF estimates. A threshold of CF of 5.0 % was used as the lower limit of detection. Participants with CF ≥ 5.0 % were classified as having mLOY, while all others were classified as not having mLOY.

### Telomere length estimation

Telomere length estimation was included as a confounder in the cross-sectional sensitivity analysis of COPDGene, with the derivation from WGS as previously described^24^. Telomere length was bioinformatically estimated using TelSeq^25^ and was batch-adjusted.

### Clonal Hematopoiesis of Indeterminate Potential (CHIP)

CHIP status was included as a confounder in the cross-sectional sensitivity analysis of COPDGene, with the derivation from WGS as previously described^26^.

### Cross-sectional analysis

Cross-sectional analyses using COPDGene data were conducted using clinical data from the same time point as the mLOY assessment, with key traits analysed including lung function measures for FEV_1_ and FEV_1_/FVC, COPD status, quantitative chest CT metrics for lung emphysema, and biological age.

### Longitudinal analysis

The longitudinal analysis included participants who attended all three phases of the COPDGene Study^13^, with a follow-up period of up ranging from 7.5 to 14.6 years. The mean follow-up time was 10.8 years. This analysis used repeated measurements collected across all three phases of the study for spirometric measures, and across Phase 1 and 2 for quantitative emphysema, and the Dunedin page of aging, to assess the association.

### Prospective analysis

A prospective analysis was conducted to evaluate whether baseline mLOY status was associated with lung function, methylation pace of aging, and incident respiratory impairment at the subsequent study phase (∼5 years later). Incident respiratory impairment included spirometric decline, increase in COPD grades and development of PRISm. If the number of incident cases (n < 15) for a given outcome was insufficient for reliable analysis, that outcome was not tested.

### TOPMed population-based cohorts

We analyzed additional population-based cohort data from the TOPMed, using WGS ‘Freeze 8’ sequencing data paired with harmonized and quality-controlled lung function and related data from the NHLBI Pooled Cohorts Study (hereafter referred to as the TOPMed population-based cohorts)^27^. Clinical data and mLOY status were matched with data from the most recent phase at the time of blood draw. Lung function measurements included pre-bronchodilator FEV_1_, FVC, FEV_1_/FVC, and COPD status as above.

To harmonize data across cohorts, several steps were taken to ensure consistency. Phenotypic variables such as smoking history, age, sex, and lung function measures were standardized by aligning units of measurement and definitions across cohorts. For example, smoking history was harmonized by ensuring consistent definitions of pack-years and smoking status, while lung function measures were checked for consistent units. Only participants with complete data were included in the analysis. Genetic ancestry PCs were generated from the WGS data.

The analysis focused on cohorts with at least 50 individuals with mLOY and spirometry, identifying five studies for cross-sectional validation: Atherosclerosis Risk in Communities (ARIC), Cardiovascular Health Study (CHS), Framingham Heart Study (FHS), Hispanic Community Health Study/Study of Latinos (HCHS/SOL), and Jackson Heart Study (JHS). For longitudinal and prospective validation, we included CHS, ARIC, FHS, and the Multi-Ethnic Study of Atherosclerosis (MESA), as these cohorts had spirometry data at multiple time points. Following CHS cohort recommendations for longitudinal and prospective analyses, only FEV_1_ was included in the multi-time point analysis. In MESA, mLOY status was assessed at baseline, whereas lung function measurements were only available at later time points (Exam 3, Exam 4, Exam 5, Exam 6, >2 years after baseline). As a result, MESA was not included in the cross-sectional analysis, which requires mLOY status and clinical data from the same time point. All participants provided written informed consent, and each study center obtained institutional review board approval.

### Statistical analysis

In COPDGene, cross-sectional associations between mLOY (used as the primary predictor) and respiratory outcomes and the epigenetic pace of aging were assessed using multivariable linear and logistic regression models, adjusting for potential confounders including age, age^2^ (to capture nonlinear age-related effects), smoking history (smoking pack-years and current smoking status), ten genetic ancestry PCs, height in centimeters, and batch PCs derived from read depth in the TOPMed WGS data^19,24^. Longitudinal measurements of lung function and epigenetic pace of aging were analysed using mixed-effects models, with baseline mLOY status as the primary predictor. These models accounted for repeated measures within participants (random effect) over time (years from baseline), with additional fixed effect covariates of age at baseline, centered age and centered age^2^ to capture age-related patterns over time. Centered age was defined as the difference between an individual’s age at the time of the visit and the median age at baseline for the entire cohort. This adjustment helps standardize age in the model and accounts for variations in age across individuals. Prospective analyses were conducted using similar multivariable models as the cross-sectional analyses, with baseline mLOY as the predictor and respiratory outcomes measured at the next follow-up visit as the outcome. Covariate adjustments included smoking status, smoking pack-years, age, age², height, genetic PCs, and batch PCs. For models assessing quantitative emphysema, scanner type was additionally included as a covariate. For the longitudinal analyses, mixed-effects model was used to account for repeated measures within participants over time (years from baseline). Fixed effects included centered age, centered age², and age at baseline. Continuous outcomes were standardized when fitting the regression models. For the association between mLOY and smoking pack-years, adjustments were made for self-reported race, smoking status, age, and age².

For the analysis in TOPMed population-based cohorts, cross-sectional, longitudinal, and prospective associations were similarly assessed, adjusting for smoking status, smoking pack-years, height, ten genetic ancestry principal components (PCs), ten batch PCs, and additionally a categorical ‘study’ variable to account for cohort-specific effects. In the cross-sectional and prospective analyses, age and age², were included as covariates.

### Sensitivity analyses

To minimize the potential confounding effect of age, we performed 1:1 nearest neighbour propensity score matching to match individuals without mLOY to those with mLOY, based on age. This matching ensured balanced age distributions between individuals with mLOY and those without in the cross-sectional sensitivity analysis.

To assess the robustness of the primary cross-sectional models in COPDGene, telomere length estimation and CHIP status were each included separately as additional covariate in sensitivity analyses.

We also performed sensitivity analyses to evaluate the robustness of our results based on variable cell fraction classification of LOY. Specifically, we reclassified individuals with mLOY based on a minimum cell fraction (CF) of 10% and repeated the analyses for both the COPDGene and TOPMed cohorts. In addition, for the prospective analysis, we included an analysis on individuals with normal lung function at baseline to further assess the stability and consistency of the association between mLOY and lung function over time.

Statistical significance was determined at *p* < 0.05 for primary analyses. All statistical analyses were performed using R (version 4.3.0).

## Results

The discovery phase involved 5,097 male participants from the COPDGene Study. Participants had self-reported ages of 40 to 85 years at the time of blood sample collection, with a smoking history of at least 10 pack-years. The median follow-up time was 5.4 years. Detailed baseline characteristics are summarized in Table 1.

**Table 1.** Descriptive statistics of the COPDGene study at baseline and follow-up time (years). Detailed trait descriptions are provided in the Methods. Abbreviations: FEV_1_, forced expiratory volume in 1 second; FVC, forced vital capacity; PRISm, preserved ratio impaired spirometry. FEV_1_/FVC, ratio of forced expiratory volume in 1 second to forced vital capacity; LAA950, percentage of low attenuation area < –950 Hounsfield units on CT (indicative of emphysema).

|  |  | without mLOY | with mLOY | *p* |
| --- | --- | --- | --- | --- |
| sample size | total (%) | 4349 (85) | 748 (15) |  |
| Cell Fractions (CF, %) | Median (IQR) | 0.0 (0.0 to 0.0) | 14.3 (9.6 to 28.9) | <0.001 |
| Age at blood draw (years) | Median (IQR) | 57.3 (51.2 to 64.5) | 68.3 (63.0 to 73.7) | <0.001 |
| Follow-up time (years) | Median (IQR) | 5.5 (0.0 to 10.2) | 5.6 (0.0 to 10.2) |  |
| Self-reported race | African American | 1661 (38.2) | 79 (10.6) | <0.001 |
|  | Non-Hispanic White | 2688 (61.8) | 669 (89.4) |  |
| Smoking status | Current smoker | 2531 (58.2) | 251 (33.6) | <0.001 |
|  | Former Smoker | 1818 (41.8) | 497 (66.4) |  |
| Sequencing center | Broad | 3488 (80.2) | 631 (84.4) | 0.009 |
|  | UW | 861 (19.8) | 117 (15.6) |  |
| Height (cm) | Median (IQR) | 176.2 (171.5 to 181.2) | 175.3 (171.3 to 180.3) | 0.001 |
| GOLD grade | Normal spirometry | 1954 (45.3) | 182 (24.5) | <0.001 |
|  | PRISm | 497 (11.5) | 51 (6.9) |  |
|  | 1 | 344 (8.0) | 89 (12.0) |  |
|  | 2 | 775 (18.0) | 204 (27.5) |  |
|  | 3 | 481 (11.2) | 141 (19.0) |  |
|  | 4 | 259 (6.0) | 75 (10.1) |  |
| Smoking pack-years | Median (IQR) | 40.2 (29.0 to 57.0) | 50.8 (37.3 to 75.0) | <0.001 |
| Post FEV<sub>1</sub> percent predicted | Median (IQR) | 81.9 (61.3 to 96.1) | 69.7 (45.0 to 87.4) | <0.001 |
| pre FEV<sub>1</sub>/FVC |  | 0.7 (0.6 to 0.8) | 0.6 (0.4 to 0.7) | <0.001 |
| post FEV<sub>1</sub>/FVC |  | 0.7 (0.6 to 0.8) | 0.6 (0.4 to 0.7) | <0.001 |
| FEV<sub>1</sub> percent predicted - GLI Global | Median (IQR) | 80.8 (60.4 to 95.5) | 72.0 (46.9 to 90.4) | <0.001 |
| FEV<sub>1</sub>/FVC percent predicted - GLI Global |  | 91.4 (76.0 to 100.0) | 79.2 (55.7 to 93.1) | <0.001 |
| LAA950 | Median (IQR) | 2.7 (0.9 to 7.3) | 5.1 (2.0 to 13.7) | <0.001 |
| log-transformed LAA 950 |  | 0.4 (-0.0 to 0.9) | 0.7 (0.3 to 1.1) | <0.001 |

We extended the analysis to additional TOPMed population-based cohorts, including ARIC, CHS, FHS, JHS, and HCHS/SOL (n = 5,345) for cross-sectional data [Supp Table 1]. Longitudinal and prospective analyses were further extended using data from ARIC, CHS, FHS, and MESA cohorts (n = 4,887 and n = 3,091, respectively) [Supp Table 2]; the median follow-up time was 5.5 years. The analysis is outlined in Figure 1.

**Figure 1.**
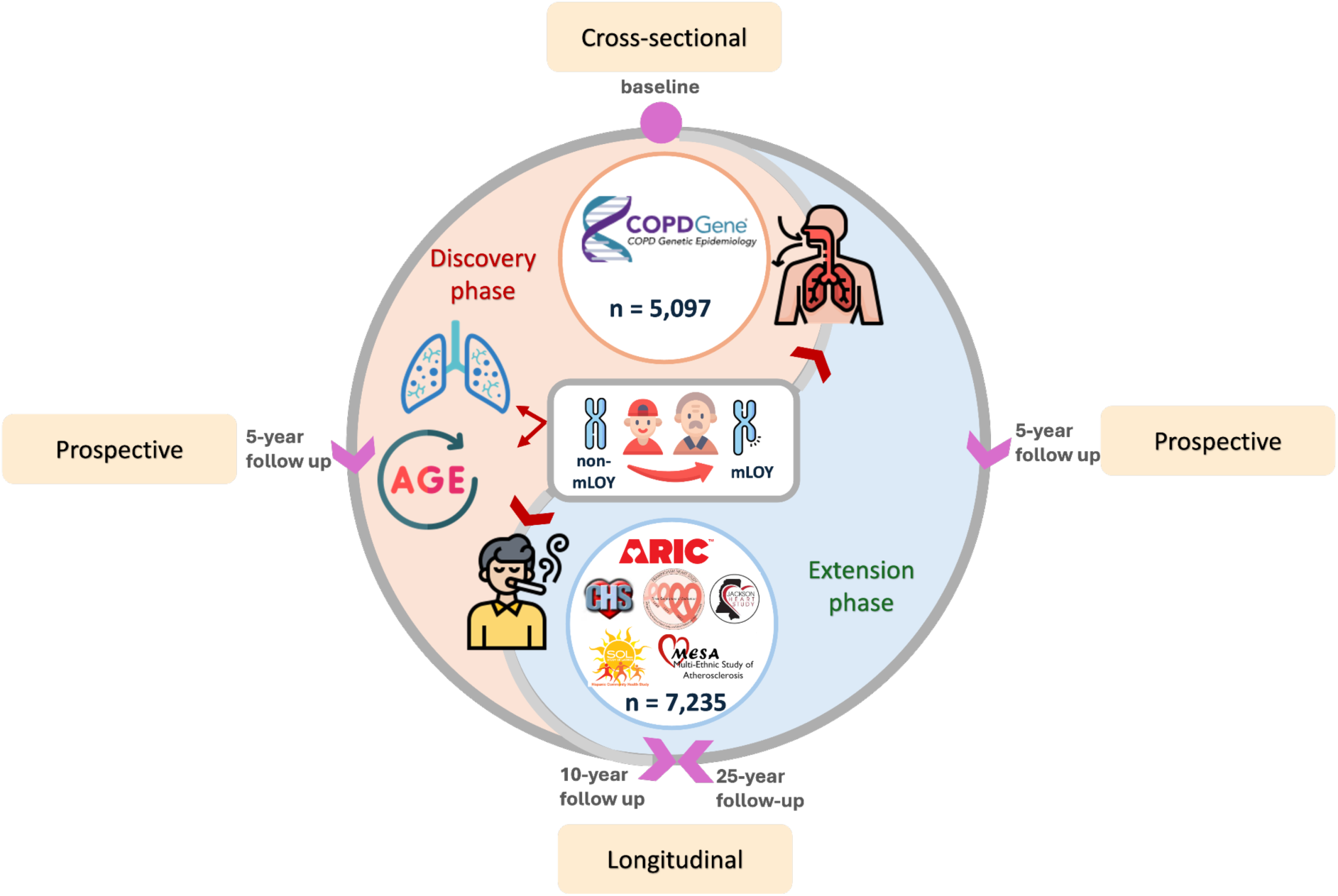
Overview of the study design, including exposure (mLOY) and outcomes (spirometry, CT scans, DNA methylation pace of aging, COPD risk) across cross-sectional, longitudinal, and prospective analyses. Discovery phase in COPDGene cohort and extension analysis using additional TOPMed population-based cohorts, including ARIC, CHS, FHS, HCHS/SOL, JHS, and MESA (see methods).

### mLOY prevalence increases with age and smoking exposure, with highest age adjusted odds in current smokers

In COPDGene, 748 individuals (14.6%) met the mLOY criteria, with a median cell fraction (CF) of 14.3% (range: 5.2% to 100%) [Table 1]. mLOY showed a positive correlation with age (r = 0.212, *p* < 5 × 10⁻⁹, Supp Figure 1A). In unadjusted analysis, former smokers had a higher prevalence of mLOY (21.48%, 497/2315) compared to current smokers (9.02%, 251/2782) (Chi-squared = 155, *p* < 2 × 10⁻¹⁶). Current smokers with mLOY were, on average, 7.49 years younger than former smokers [Supp Figure 1B]. After adjusting for age, age², smoking status and race, current smokers had 1.25-fold higher odds of mLOY than former smokers (*p* = 0.026), and mLOY prevalence increased with pack-years of cigarettes smoked (OR = 1.004 per pack-year, *p* = 0.012) [Supp Figure 1C].

In the TOPMed cohorts, 819 individuals (15.3%) had mLOY, with a median CF of 14.0% (range: 5.1% to 68.0%) [Supp Table 1]. Like the COPDGene cohort, mLOY increased with age (r = 0.282, *p* < 2 × 10⁻¹⁶, Supp Figure 1D). In unadjusted analysis, former smokers had a higher mLOY prevalence (20%, 463/2316) than current smokers (14.9%, 157/1055) or never smokers (9.98%, 195/1953) [Supp Table 1]. Current smokers with mLOY were ∼8 years younger than former and never smokers, respectively (*p* < 0.001) [Supp Figure 1E]. After age adjustment, current smokers had 2.03-fold and 2.75-fold higher odds of mLOY compared to former and never smokers, respectively (*p* = 2.30 × 10⁻⁹, *p* = 1.81 × 10⁻¹⁴). mLOY prevalence was positively associated with smoking pack-years (OR = 1.005 per pack-year, *p* = 0.009) after adjusting for smoking status, race, study, age, and age² [Supp Figure 1F].

### mLOY is associated with lower lung function, higher emphysema, and faster pace of epigenetic aging

In COPDGene, individuals with mLOY exhibited significantly lower lung function compared to those without mLOY. This difference was observed for both pre- and post-bronchodilator FEV_1_/FVC. Specifically, pre-FEV_1_/FVC was reduced by −0.115 (95% CI: −0.190 to −0.040, *p* = 2.65 × 10⁻³), and post-FEV_1_/FVC showed a similar decrease of −0.122 (95% CI: −0.196 to −0.048, *p* = 1.24 × 10⁻³) [Figure 2, Supp Figure 2]. mLOY showed a suggestive association with higher prevalence of COPD (*p* = 0.06) [Figure 2].

**Figure 2.**
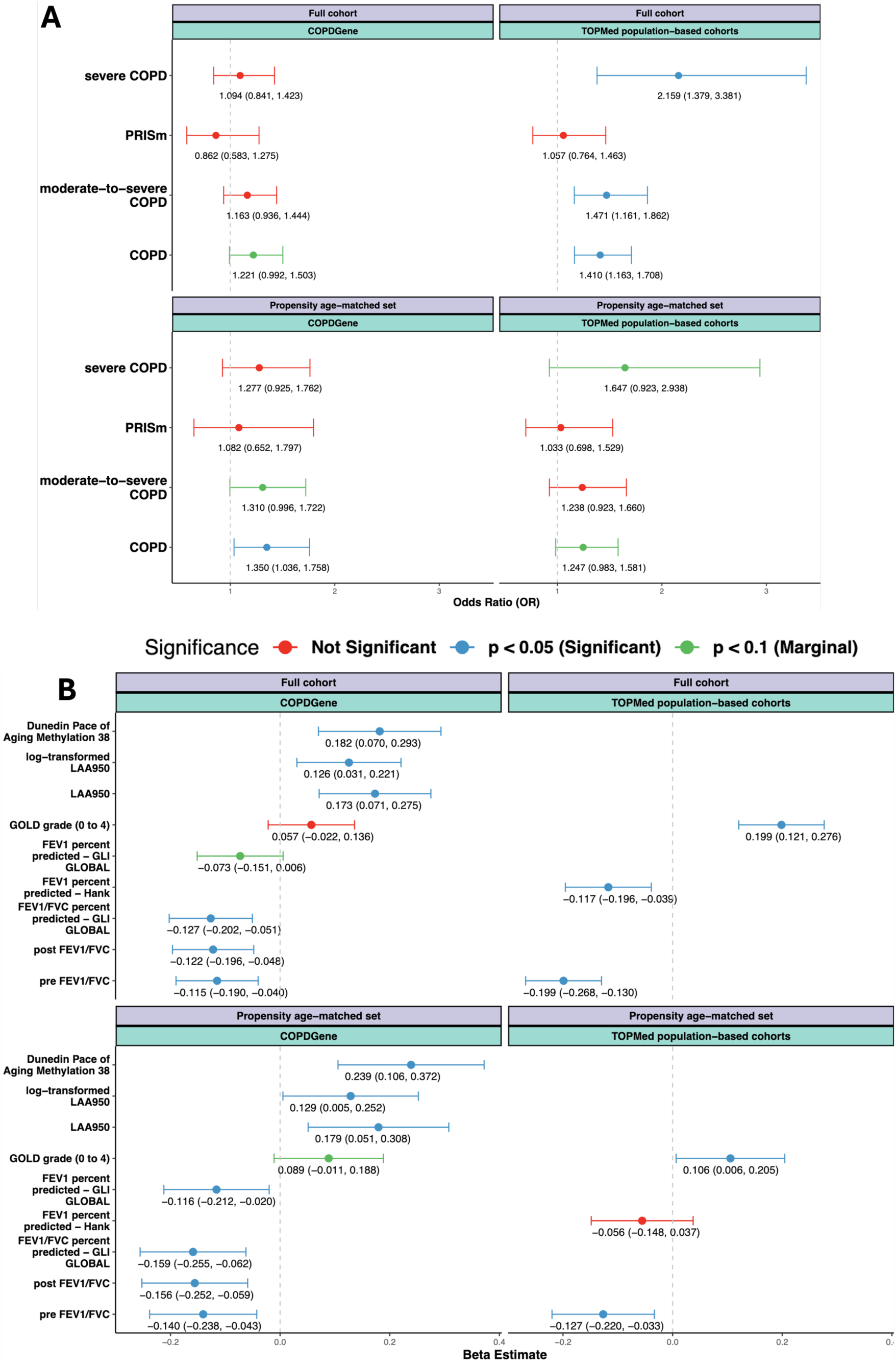
Forest plots showing (A) odds ratios for binary traits and (B) beta coefficients for continuous traits, with 95% confidence intervals (CI), in COPDGene and TOPMed population-based cohorts. Analyses are presented for both the full cohort and an age-matched propensity score subset, with adjustments for ten genetic ancestry principal components (PCs) and additional confounders as detailed in the results section. Statistical significance is color-coded.

Similar associations were observed in the TOPMed cohorts, where individuals with mLOY also demonstrated significantly lower FEV_1_/FVC (−0.199, 95% CI: −0.268 to −0.130, *p* = 1.84 × 10^-8^), lower FEV_1_ (−0.117, 95%CI: −0.196 to −0.039, *p =* 0.003)and higher GOLD grade (0.199, 95% CI: 0.121 to 0.276, *p* = 6.02 × 10^-7^) [Figure 2]. Following this, we repeated analyses stratified by study, and the relationship between mLOY and lower lung function remained significant across each individual study cohort [Supp Figure 3, Supp Table 3]. mLOY was also significantly associated with COPD (overall, moderate-to-severe COPD, and severe COPD [Figure 2]).

**Figure 3.**
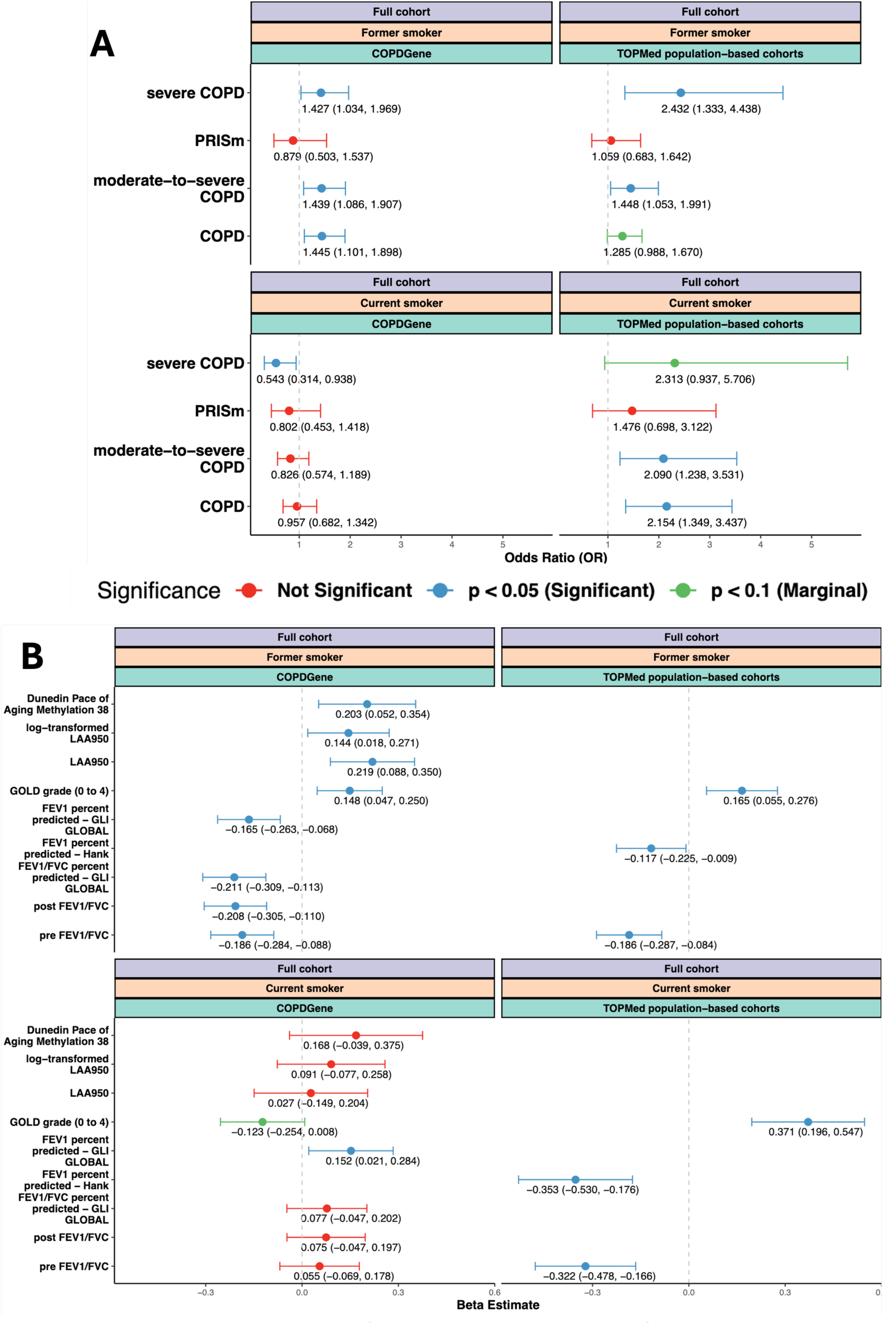
Forest plots showing (A) odds ratios for binary traits and (B) beta coefficients for continuous traits, with 95% confidence intervals (CIs), in COPDGene and TOPMed population-based cohorts, stratified by smoking status. Results for never smokers in the TOPMed cohort are shown in Supplementary Figure 5B. All models are adjusted for ten genetic ancestry principal components (PCs) and additional confounders as described in the Results section. Statistical significance is color-coded.

In COPDGene, we also found that mLOY was positively associated with quantitative emphysema (measured by log-transformed LAA950), showing an effect size of 0.126 (95% CI: 0.031 to 0.221, *p* = 9.62 × 10⁻³) and with accelerated epigenetic pace of aging, with an effect size of 0.182 (95% CI: 0.070 to 0.293, *p* = 1.46 × 10⁻³) [Figure 2, Supp Figure 2].

Age-matched propensity analyses confirmed that mLOY was significantly associated with lower lung function in both COPDGene and TOPMed, and with increased emphysema and accelerated epigenetic aging in COPDGene [Figure 2, Supp Table 4-5].

### Interaction between mLOY and smoking

We tested whether pack-years modified the association between mLOY and respiratory outcomes or epigenetic aging using interaction models. mLOY was strongly associated with epigenetic aging (effect size = 0.476, 95% CI 0.243-0.709, *p* = 6.27 x 10⁻^5^), and smoking pack-years showed a direct effect size per pack-year (effect size = 0.007, 95% CI 0.005-0.008, *p* = 1.06 × 10⁻¹²). A significant interaction between mLOY and pack-years was observed (interaction effect size = −0.006, 95% CI −0.009 to −0.002, *p* = 0.005) [Supp Figure 4A]. The mLOY effect on epigenetic aging was most evident in the 30-41.3 pack-year group (*p* = 6.36 x 10^-5^) [Supp Figure 4B].

**Figure 4.**
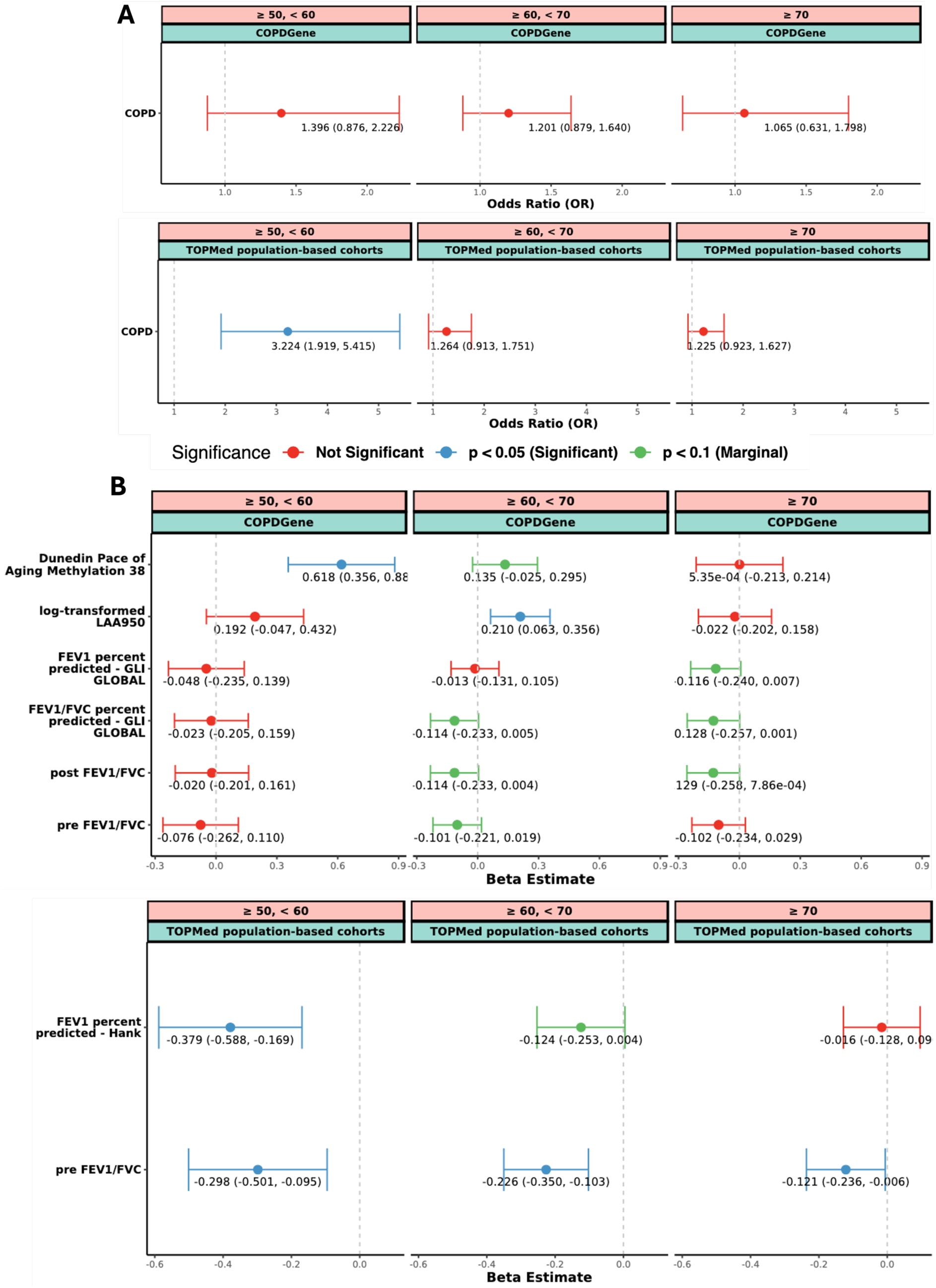
Forest plots displaying beta coefficients with 95% confidence intervals (CIs) from age-stratified analyses in the COPDGene and TOPMed population-based cohorts. Models are adjusted for ten genetic ancestry principal components (PCs) and additional confounders as described in the Results section. Statistical significance is color-coded.

We also explored the interaction between mLOY and smoking status on lung function. A significant interaction was observed, with the negative effect of mLOY on lung function more pronounced in former smokers than in current smokers, with former smokers showing a lower post-bronchodilator FEV_1_/FVC (effect size = −0.202, 95% CI −0.348 to −0.056, *p* = 0.007) compared to current smokers [Supp Figure 4C].

This prompted a stratified analysis by smoking status. Among former smokers (median age: 65.6), mLOY was significantly associated with COPD risk (COPD (OR = 1.445), moderate-to-severe COPD (OR = 1.439), and severe COPD (OR = 1.427)) [Figure 3, Supp Table 6]. Former smokers with mLOY also exhibited significantly lower lung function (post-bronchodilator FEV_1_/FVC; effect size = −0.208, 95% CI: −0.305 to −0.110, *p* = 2.98 × 10⁻⁵), greater emphysema (log-transformed LAA950; effect size = 0.144, 95% CI: 0.018 to 0.271, *p* = 0.026), and faster epigenetic pace of aging (effect size = 0.203, 95% CI: 0.052 to 0.354, *p* = 0.009) [Figure 3, Supp Table 6]. In contrast, associations were attenuated in current smokers (median age: 54.1) [Figure 3, Supp Table 6]. Given the substantial age difference between the groups, we performed age propensity-matched stratified analyses to reduce confounding. The associations with mLOY remained significant in former smokers, while no significant associations were observed in current smokers [Supp Figure 5, Supp Table 7].

**Figure 5.**
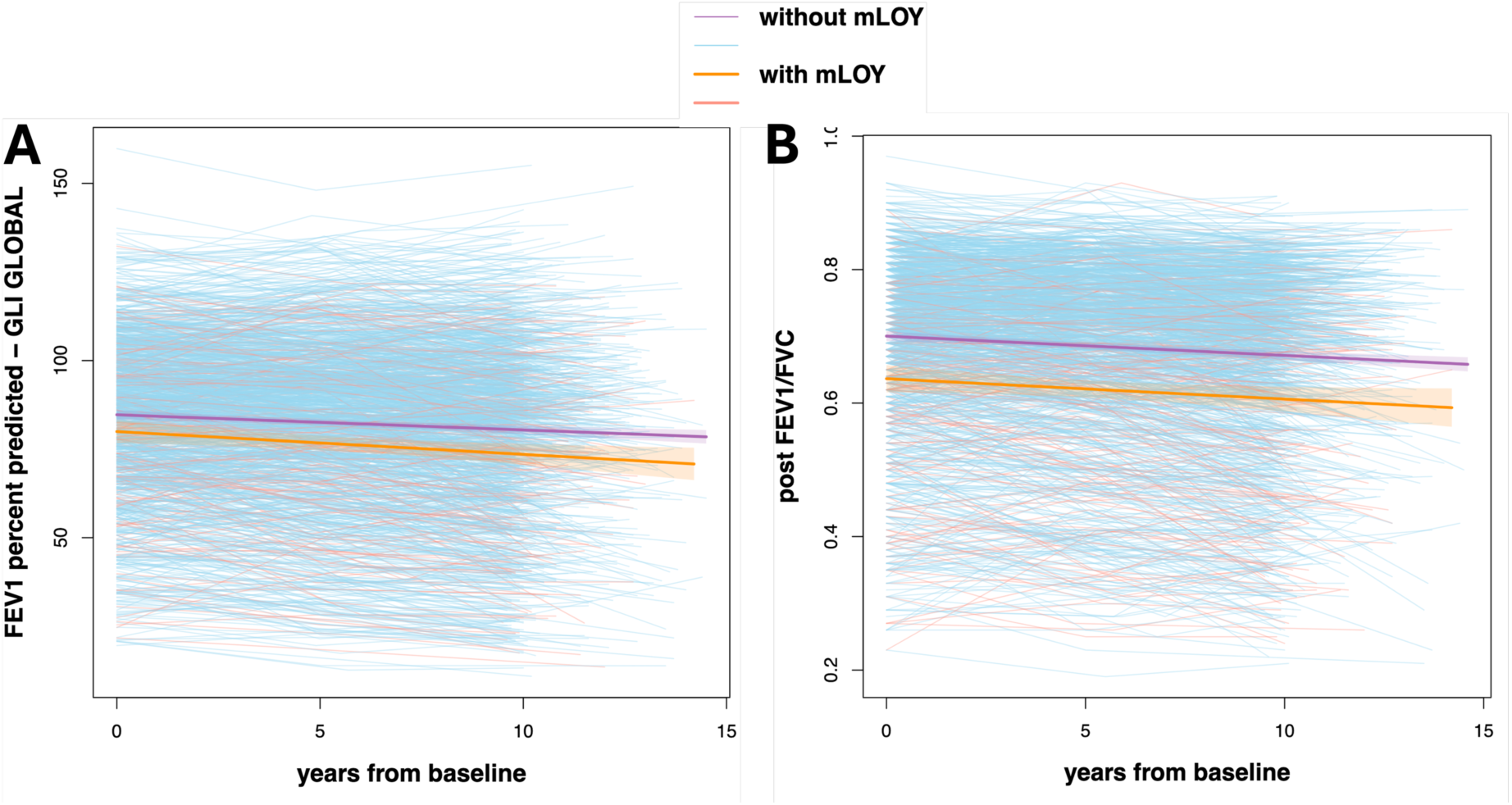
Scatter plots showing the distribution of (A) FEV₁ percent predicted (using the GLI GLOBAL equation; see Methods) and (B) post-bronchodilator FEV₁/FVC ratios with regression coefficients and 95% confidence intervals (CIs), for individuals who participated in all study phases (Phase 1 to 3) of the longitudinal analysis. The corresponding forest plot is available in Supplementary Figure 7. Individual lines are color-coded by mLOY status (with vs. without mLOY).

In the TOPMed population-based cohorts, in interaction models with smoking status, mLOY remained significantly associated with lower lung function [Supp Figure 6A, Supp Table 8]. Importantly, stratified analyses showed that mLOY was associated with lower lung function in both former (median age: 65) and current (median age: 54) smokers [Figure 3, Supp Table 8], but not in never-smokers [Supp Figure 6B, Supp Table 8]. The associations for FEV_1_/FVC remained significant in current smokers after age-propensity matching, were attenuated in former smokers, and were not significant in never smokers both before and after matching [Supp Figure 5, Supp Table 9]. The associations for FEV_1_ percent predicted remained significant in current smokers [Supp Figure 5, Supp Table 9].

**Figure 6.**
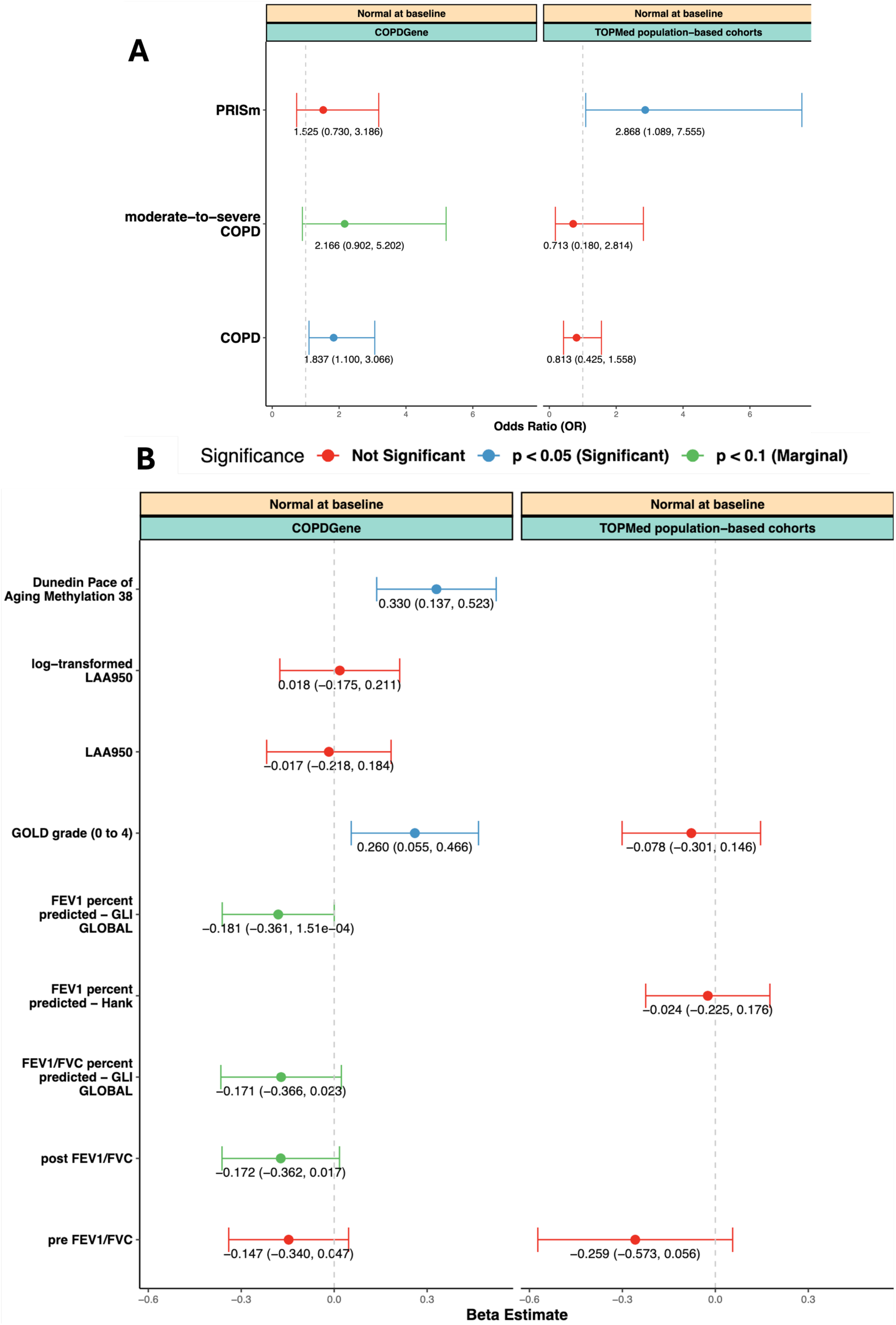
Forest plots displaying beta coefficients for continuous traits with 95% confidence intervals (CIs) from the prospective analysis of the COPDGene and TOPMed population-based cohorts. The analysis focuses on individuals with normal lung function at baseline, as well as those maintaining normal lung function at both baseline and follow-up. Adjustments are made for ten genetic ancestry principal components (PCs) and additional confounders as outlined in the Results section. Statistical significance is color-coded.

### mLOY showed age-dependent associations with lung function, emphysema and aging in COPDGene, but no age-dependency in additional TOPMed population-based cohorts

The interaction between mLOY and age was statistically significant for epigenetic pace of aging. Age stratification was conducted to better assess the potential variation in mLOY’s association with lung function across different age groups. Individuals under 50 were excluded due to the lower prevalence of mLOY in this age group (as noted in Jakubek et al^23^). The remaining participants were divided as described in Jakubek et al^23^ into three age groups, 50 to 60 years old (n = 1,872), 60 to 70 years old (n = 1,546), and more than 70 years old (n = 824).

In COPDGene, despite the lower numbers in the age strata [Supp Table 10], mLOY demonstrated trends for association with lower lung function, particularly in individuals over 60 (≥ 70 age group: −0.129, p = 0.052; 60 to 70 age group: −0.114, *p* = 0.059) [Figure 4, Supp Table 11]. mLOY was also associated with faster epigenetic pace of aging in the 50-60 age group (effect size = 0.618, *p* = 4.61 × 10⁻⁶) and increased emphysema in the 60-70 age group (effect size = 0.210, *p* = 0.005) [Figure 4, Supp Table 11].

In the TOPMed population-based cohorts, mLOY showed a consistent association with lower lung function across all age groups and a positive association with GOLD grade [Figure 4, Supp Table 12-13]. These findings suggest that mLOY is consistently associated with impaired lung function, with variations in its effects depending on age and study cohort exposure histories.

### COPDGene shows mLOY association with lung function decline

A longitudinal analysis in COPDGene including 1,776 individuals (follow-up time median: 10.4 years, mean 10.8 years) showed that mLOY was associated with a significant decline in FEV_1_ percent predicted by GLI over time (effect size = −0.022, *p* = 4.35 x 10^-4^) and a trend toward FEV_1_/FVC decline (effect size = −0.010, *p* = 0.101), and [Figure 5, Supp Figure 7]. However, when we stratified by smoking status, both former and current smokers showed the time-by-mLOY was not significant, although the direction was consistent [Supp Figure 7]. In the TOPMed cohorts, no significant longitudinal associations between mLOY and lung function decline were observed across the four contributing cohorts.

### Baseline mLOY is associated with accelerated epigenetic aging and lower lung function in individuals with normal spirometry

In COPDGene, analysis of 1,373 individuals with normal spirometry at baseline suggests that mLOY may be an early indicator of epigenetic biological aging. Baseline mLOY was significantly associated with accelerated pace of epigenetic aging (effect size = 0.330, 95% CI: 0.137 to 0.522, *p* = 8.25 × 10⁻⁴) [Figure 6, Supp Table 14]. We observed an increased risk of COPD (OR = 1.84, 95% CI 1.10, 3.07, *p* = 0.02) in individuals with normal lung function at baseline [Figure 6, Supp Table 14]. A marginal association was also observed between baseline mLOY and both lower future FEV_1_ GLI and FEV_1_/FVC [Figure 6, Supp Table 14].

Among individuals with normal lung function at baseline (n = 723) in TOPMed population-based cohorts, baseline mLOY was significantly associated with increased risk of progression to PRISm (OR = 2.868, 95% CI: 1.089 to 7.555, *p* = 0.033) [Figure 6, Supp Table 15].

In the full COPDGene prospective cohort (n = 3,070), baseline mLOY was associated with subsequent visit lower pre-and post-bronchodilator FEV₁/FVC (Pre-, −0.177; 95% CI: −0.283 to −0.072; *p* = 0.001; post-, −0.186, 95% CI −0.291 to −0.082, *p* = 4.95 x 10⁻^4^) and accelerated epigenetic aging (0.180; 95% CI: 0.071 to 0.290; *p* = 0.001) [Supp Figure 8]. After adjusting for baseline spirometry, the significance of these associations attenuated, although the direction of effects remained similar.

### Robustness of mLOY associations at more stringent cell fraction thresholds and with aging-related covariates

In all main analyses, individuals with mLOY were classified using a CF threshold of ≥5%. As a sensitivity analysis, we also used a higher threshold of ≥10% in both COPDGene (n = 535) and TOPMed (n = 552). Despite fewer mLOY cases, the association with key outcomes remained consistent, reinforcing result robustness. Effect sizes were slightly stronger at CF ≥10%, suggesting a possible dose-dependent relationship [Supp Figure 9, Supp Table 16-17].

In COPDGene cross-sectional models, inclusion of CHIP status as a covariate yielded consistent mLOY associations with slightly stronger effect sizes [Supp Figure 10, Supp Table 18]. Similarly, adjustment for telomere length retained the mLOY association, with only modest attenuation of effect sizes (∼0.01) [Supp Figure 10, Supp Table 18].

## Discussion

Our study leveraged measured spirometry, CT emphysema, epigenetic biological aging from COPDGene and also measured spirometry from six additional TOPMed cohorts (ARIC, CHS, FHS, HCHS/SOL, JHS, and MESA) to explore the association between mLOY and respiratory outcomes. Across both cross-sectional and prospective analyses, mLOY emerged as a robust marker of impaired lung function. In COPDGene, mLOY was significantly associated with lower lung function (FEV_1_/FVC), increased emphysema, and accelerated epigenetic aging. The FEV_1_/FVC and FEV_1_ associations were similarly observed in the additional TOPMed cohorts, strengthening the generalizability of this relationship. Longitudinally, mLOY was associated with accelerated lung function decline. Among individuals with normal lung function at baseline, mLOY showed a trend toward lower lung function and significant accelerated epigenetic aging in COPDGene, and the association with PRISm incidence in the TOPMed cohorts. Epigenetic pace of aging was strongly associated with mLOY, further reinforcing its role in lung aging. The associations remained robust after adjusting for aging-related factors such as telomere length and CHIP. Overall, these findings support mLOY as a biomarker for lung function impairment, especially in individuals who have already stopped smoking.

Both the COPDGene and TOPMed cohorts demonstrate an age-related increase in mLOY prevalence, highlighting its role as a marker of cellular aging in some men, which may contribute to an underlying genetic predisposition to aging-related phenotypes or diseases. Recent findings have associated mLOY with increased risks of COPD (classified by ICD 10 codes), lung cancer and idiopathic pulmonary fibrosis^10–12^. The observed association between mLOY and lower lung function suggests that mLOY as a somatic mutation may be an early marker of accelerated epigenetic aging processes within the lung, potentially elevating the risk of lung function impairment over time. As mLOY reflects chromosomal loss linked to cellular senescence, its influence on lung health may parallel broader aging mechanisms affecting other tissues and organ systems^28^.

Smoking is a well-established risk factor for lung function decline^29^, and the combination of smoking with mLOY may enhance smoking-induced damage^8,30,31^. Among the COPDGene participants who previously smoked, mLOY was significantly associated with a higher risk of COPD. They may have already endured prolonged periods of smoking damage and may exhibit more severe consequences associated with mLOY due to the compounded interacting effects of genetic predisposition, somatic mutation, aging and cumulative environmental exposures. This is a critical observation as many studies to date^4,7,9,10^ associate mLOY with current smoking. The findings suggest that mLOY may accelerate the harmful effects of smoking on lung function, particularly in individuals with a long smoking history and higher intensity of smoking, even after smoking cessation or adjusted for CHIP status or telomere length.

The COPDGene cohort highlights associations between mLOY and lung function across different age groups. In younger adults (50 to 60 years), mLOY is strongly associated with accelerated epigenetic aging, which, combined with smoking, may contribute to earlier manifestations of pulmonary dysfunction, including declining lung function, symptoms and emphysema. In adults aged 60 to 70 years, mLOY is marginally associated with increased emphysema, suggesting a cumulative effect with smoking. In those over 70, mLOY is marginally associated with lower lung function, raising the possibility that cumulative effects of somatic mutation, aging, and smoking may contribute to lung impairment. These findings highlight the importance of considering both the timing of the emergence of mLOY and smoking history when evaluating long-term respiratory outcomes and supports the need for large scale longitudinal studies.

The relationship between mLOY and lung function varies across cohorts and study designs, possibly due to lower smoking exposure in some populations. Longitudinal analysis in COPDGene shows FEV_1_ decline with mLOY, while TOPMed cohorts show no significant time interactions, possibly due to differences in cohort composition or smoking effects. Prospective analysis in both cohorts supports mLOY as an early indicator of future lung function decline, highlighting its potential as a clinically translatable biomarker for lung health and COPD severity.

Individuals with normal lung function at the time of mLOY measurement may show subtle lung function impairment over time, as seen in COPDGene. Similar findings in TOPMed cohorts support mLOY as a generalizable indicator of lung disease risk. This highlights the importance of monitoring smokers with spirometry and biomarkers like mLOY. The association between mLOY and accelerated epigenetic aging suggests early cellular changes that could precede COPD. These results highlight the need for further research into mLOY’s long-term effects and potential causal mechanisms, especially in individuals at risk of disease progression. Supporting this, Regan et al^32^ demonstrated that smokers without spirometric evidence of COPD may still have significant respiratory impairments, such as physical function decline, worse quality of life, and CT evidence of emphysema. Regan et al^32^ emphasized that relying solely on spirometry may miss underlying lung damage, particularly in individuals over 55, stressing the need for broader diagnostic approaches, including biomarkers, imaging and symptom assessment, to identify at-risk individuals for early intervention^28^.

The strength of our study lies in its comprehensive and granular analysis of mLOY and its association with lung health, uniquely integrating careful phenotyping including spirometry measures, quantitative emphysema, and biological aging across cross-sectional, longitudinal, and prospective analyses, in multi-ancestry cohorts with variable histories of cigarette smoking. We provide a detailed examination of how varying levels of mLOY burden relate to early lung function impairment and disease progression, highlighting its potential as an pre-clinical biomarker for respiratory decline. Findings from the additional TOPMed population-based cohorts further emphasizes the robustness and generalizability of mLOY as a potential predictive marker for respiratory health deterioration. Limitations include potential residual confounding and burden of multiple testing; however, findings remained robust across multiple cohorts and sensitivity analyses. The differences observed between cross-sectional and longitudinal findings highlight the strengths of longitudinal analyses in capturing progressive effects of mLOY. Longitudinal trends suggest that mLOY is associated with adverse lung function outcomes over time, reinforcing the value of longitudinal designs for uncovering progressive effects, and the need for larger and longer prospective studies to fully evaluate mLOY’s role in lung aging and decline.

In conclusion, our study provides compelling evidence that mLOY is associated with COPD, lower lung function, increased emphysema, and accelerated epigenetic aging. These findings highlight the potential of mLOY as a clinical biomarker for progressive lung function decline and emphasizes the importance of considering both somatic mutations and environmental factors in managing respiratory health. Further research is needed to understand the underlying mechanisms driving these associations and the implications for targeted interventions for men at risk for COPD.

## Data Availability

All data produced are available online at https://dbgap.ncbi.nlm.nih.gov/beta/home.
ARIC(phs001211), CHS (phs001368), COPDGene(phs000951), FHS(phs000974), HCHS/SOL(phs001395), JHS(phs000964), and MESA(phs001416 )

## Acknowledgements

We would like to thank all the participants in the COPDGene and TOPMed population-based cohorts (including ARIC, CHS, FHS, HCHS/SOL, JHS, and MESA) for their participation. We also acknowledge the financial support provided by K24 HL171900, R01 HG011393, and 75N92023D00011. E.C.O is supported by R21HL165405, R21HL153700, K23HL130627, R21HL129924. A.M is supported by R01HL153248. Molecular data for the Trans-Omics in Precision Medicine (TOPMed) program was supported by the National Heart, Lung, and Blood Institute (NHLBI). The views expressed in this manuscript are those of the authors and do not necessarily represent the views of the NHLBI, the National Institutes of Health, or the US Department of Health and Human Services. The full study acknowledgements are detailed in the supplemental information as extended acknowledgements.

