## Supplementary Material for "Mosaic Loss of Y chromosome associates with lung function, emphysema and epigenetic aging"

### Supplementary Information

### Cohort Information of six NHLBI cohort studies

The extension phase in this study included 7,235 individuals from six NHLBI TOPMed studies: ARIC (n = 1,295), CHS (n = 1,093), FHS (n = 687), HCHS/SOL (n = 1,357), JHS (n = 913), and MESA (n = 1,890). Cross-sectional was conducted using data from five cohorts: ARIC, CHS, FHS, JHS, and HCHS/SOL, comprising 5,345 participants. Descriptive summary statistics for the additional TOPMed population-based cross-sectional cohorts are provided in Supplementary Table 1. Longitudinal included data from ARIC, CHS, FHS, and MESA (n = 4,887), with corresponding descriptive statistics presented in Supplementary Table 2. Prospective was performed using a subset of the same four cohorts (n = 3,091). All participants provided written informed consent across the contributing studies[1-8].

*ARIC*

The Atherosclerosis Risk in Communities (ARIC)[1,2] study is a population-based prospective cohort study of cardiovascular disease, initiated in 1985 and sponsored by the National Heart, Lung, and Blood Institute (NHLBI). The study was designed to monitor trends in myocardial infarction (MI) incidence and coronary heart disease (CHD) mortality across four U.S. communities: Forsyth County, North Carolina; Jackson, Mississippi; suburban Minneapolis, Minnesota; and Washington County, Maryland. Between 1987 and 1989, ARIC enrolled 15,792 predominantly African American and European American adults aged 45 to 64 years, selected through probability sampling from these communities. Since baseline, participants have been followed for more than three decades, with extensive data collection through phone interviews and in-person clinic visits. To date, ten examination visits have been conducted, including triennial follow-up exams and more recent exams in 2011–2013 (Visit 5), 2016–2017 (Visit 6), 2018–2019 (Visit 7), 2020 (Visit 8), 2021–2022 (Visit 9), and 2023 (Visit 10). In addition to cohort follow-up, ARIC conducted active community surveillance of cardiovascular events from 1987 through 2014, providing annual estimates of acute MI incidence, CHD mortality, and case fatality rates in the four study regions. Over time, the study has adopted novel data collection technologies, including wearable ECG patches and comprehensive cognitive assessments, to support research into vascular contributions to cognitive impairment and dementia (VCID). With more than 2,300 peer-reviewed publications, ARIC has made substantial contributions to cardiovascular and aging research. The study's inception was influenced by the 1978 NHLBI Conference on the Decline in CHD Mortality, which emphasized the need for longitudinal studies to investigate the causes of declining heart disease mortality observed in the previous decade. The ARIC study has since become a cornerstone of epidemiologic research in cardiovascular health and disease.

*CHS*

The Cardiovascular Health Study (CHS)[3] is a population-based, longitudinal cohort study of coronary heart disease and stroke in adults aged 65 years and older. The study aims to identify both traditional and novel risk factors for cardiovascular disease (CVD) in older adults, with an emphasis on modifiable and protective factors. CHS enrolled 5,201 predominantly European American participants in 1989–1990, selected from Medicare eligibility lists in four U.S. communities: Forsyth County, NC; Sacramento County, CA; Washington County, MD; and Pittsburgh, PA. In 1992–1993, an additional 687 predominantly African American participants were enrolled, bringing the total cohort to 5,888. At baseline, participants underwent extensive clinical and laboratory evaluations to assess CVD risk factors, subclinical disease, and prevalent CVD. Annual examinations continued through 1999, followed by ongoing surveillance for stroke and cardiovascular events. Blood samples were collected at baseline, and DNA was extracted from participants who consented to genetic studies.

*FHS*

The Framingham Heart Study (FHS)[4] is a long-running, multi-generational, community-based research project aimed at understanding the risk factors and natural progression of cardiovascular disease (CVD), including stroke. The study began in 1948, with the Original Cohort consisting of 5,209 adults from Framingham, Massachusetts, two-thirds of the town's adult population at the time. Over the decades, the study has expanded to include two additional cohorts: the Offspring Cohort, recruited in 1971, and the Third Generation Cohort, enrolled starting in 2002. The Original Cohort participants, who were followed closely since 1948, continue to undergo biennial exams to track long-term health outcomes. The Offspring Cohort includes 5,124 individuals (with 3,514 being biological children of the Original Cohort), and they have been examined approximately once every 4 years. The Third Generation Cohort comprises 4,095 participants, all of whom are at least 20 years old and have at least one parent from the Offspring Cohort. The study is comprehensive in its design and data collection, incorporating both phenotypic (observable traits) and genotypic (genetic) information from the different generations. The goal is to understand the genetic and environmental factors influencing cardiovascular, lung diseases, and other health conditions. The Third Generation Cohort was specifically designed to complement the data gathered from the Original and Offspring Cohorts, with particular emphasis on larger families for genetic studies. This cohort was recruited through extensive outreach to potential participants, including direct invitations to eligible individuals, with priority given to those from large extended families that span three generations. Since 2002, participants in the Third Generation Cohort have been assessed with clinical and laboratory examinations, focusing on vascular risk factors and subclinical atherosclerosis, alongside measures of cardiac structure and function. This data is crucial for exploring how cardiovascular conditions develop over time, the genetic predispositions involved, and the role of environmental influences. By comparing these newer data points to the older data from the Original and Offspring Cohorts, the study aims to uncover important genetic and environmental determinants of CVD, stroke, and lung disorders, with a particular focus on cardiovascular diseases. In total, the Framingham Heart Study spans three generations, with multi-decade follow-ups and ongoing examinations to monitor the health trajectories of participants. Over its nearly 60 years of research, it has been instrumental in identifying key risk factors for cardiovascular diseases and other health conditions, and its data continues to inform public health strategies and clinical practices related to cardiovascular health.

*JHS*

The Jackson Heart Study (JHS)[5,6] is a large, community-based, longitudinal cohort study designed to investigate the biological, behavioral, and psychosocial determinants of cardiovascular disease (CVD) among African American adults. Launched in 2000, JHS is the largest single-site epidemiologic study of CVD in African Americans and aims not only to explore risk factors for atherosclerotic and cardiometabolic outcomes, but also to enhance the participation of African American communities and scientists in biomedical research. The cohort consists of 5,306 non-institutionalized African American adults aged 35 to 84 years at enrollment, drawn from urban and rural areas of the Jackson, Mississippi metropolitan statistical area (Hinds, Madison, and Rankin counties). Participants were recruited through a combination of methods: random sampling (17%), volunteers (30%), participants from the Atherosclerosis Risk in Communities (ARIC) Study (31%), and family members of enrolled participants (22%). A subset of younger adults (ages 21–34) was also enrolled in a nested family cohort. Baseline data collection (Exam 1: 2000–2004) included detailed medical history, physical examination, laboratory tests, and psychosocial and lifestyle questionnaires addressing factors such as physical activity, spirituality, coping strategies, perceived discrimination, socioeconomic status, and healthcare access. Follow-up clinic visits occurred in 2005–2008 (Exam 2) and 2009–2013 (Exam 3), with over 80% of surviving participants retained across exams. In addition to clinic visits, annual follow-up calls have been conducted since baseline to update medical histories, assess functional status, and document hospitalizations and clinical events. The study includes extensive clinical phenotyping such as electrocardiography, echocardiography, carotid ultrasound, ankle-brachial index, and CT and MRI imaging for vascular and fat distribution assessments. Ongoing surveillance is conducted via medical record abstraction, linkage to CMS data, and adjudication of cardiovascular events and mortality. Genetic data, including whole genome sequencing for over 3,400 participants, have been deposited in dbGaP for public use. The JHS also maintains an active community outreach and education program, alongside public health training initiatives aimed at fostering diversity in biomedical research.

*HCHS/SOL*

The Hispanic Community Health Study/Study of Latinos (HCHS/SOL)[7] is a large, multi-center cohort study designed to explore the role of acculturation and other factors influencing the health of Hispanic/Latino populations in the U.S. Sponsored by the National Heart, Lung, and Blood Institute (NHLBI) and other NIH institutes, the study began in 2006 with a target population of 16,000 self-identified Hispanic/Latino individuals, aged 18-74. Participants were recruited from four U.S. cities, including San Diego, CA; Chicago, IL; Miami, FL; and the Bronx, NY, which represented a range of Latino ethnic backgrounds, including Cuban, Puerto Rican, Dominican, Mexican, and Central/South American origins. Recruitment was conducted using a two-stage area household probability design, and extensive psycho-social and clinical assessments, including baseline and post-albuterol challenge spirometry, took place from 2008 to 2011. Participants have been followed annually through telephone interviews since the study's initiation, with re-examinations held during a second visit from

2014 to 2017 and a third visit between 2020 and 2024. The primary goals of the study include investigating the prevalence and development of heart and lung diseases, along with other chronic conditions such as diabetes, obesity, and asthma.

*MESA*

The Multi-Ethnic Study of Atherosclerosis (MESA)[8] is an extensive research project designed to investigate the characteristics and risk factors of subclinical cardiovascular disease (CVD), a form of cardiovascular disease that is detected before clinical symptoms appear, and to track its progression to clinically overt CVD. The study focuses on understanding how various risk factors contribute to both the development and progression of subclinical cardiovascular disease. Launched in 2000, MESA initially enrolled 6,814 asymptomatic men and women between the ages of 45 and 84, all free from known cardiovascular disease at the start. The participant pool is ethnically diverse, consisting of 38% White, 28% African American, 22% Hispanic, and 12% Chinese individuals, ensuring broad representation across various race and ethnicity groups. These participants were recruited from six field centers across the United States: Wake Forest University (North Carolina), Columbia University (New York), Johns Hopkins University (Maryland), University of Minnesota (Minnesota), Northwestern University (Illinois), and University of California - Los Angeles (California). Blood samples were taken from participants to assess biochemical risk factors, and DNA samples were collected. The study design originally included 4 follow-up visits over 7 years, later expanded to 6 follow-up visits, tracking participants for over 25 years. These visits were used to monitor progression of subclinical atherosclerosis, evaluate the stability of risk factors, and collect new health measures. MESA participants were also regularly contacted every 9 to 12 months for updates on clinical events, including acute myocardial infarction, angina, stroke, congestive heart failure (CHF), and peripheral vascular disease. Mortality data and treatment information, such as revascularization procedures, were also systematically collected and analyzed.

*Smoking History in additional TOPMed population-based cohorts*

Some participants in the pooled cohort data have inconsistencies in smoking history data, particularly the former smokers with 0 pack-years listed (n = 46 in ARIC; n = 1 in CHS; n = 20 in FHS). We conducted several sensitivity analyses to assess the robustness of our findings. First, we examined the effect of removing these data points by excluding individuals with 0 pack-years from the analysis. Additionally, we explored the impact of removing smoking-related covariates, such as smoking pack-years, smoking status and cigarettes smoked per day, from the models. These sensitivity analyses were performed to determine whether the inclusion or exclusion of these participants significantly affected the results. The results of these sensitivity analyses did not substantially differ from the original models, suggesting that the inclusion of former smokers with 0 pack-years did not notably alter the association between mLOY and lung function, or COPD.

*Note on Smoking Status in HCHS/SOL*

In our extension analysis using additional TOPMed population-based cohorts, smoking status was based on participants' self-reported classification as "current smokers." However, within the HCHS/SOL cohort, a significant proportion of individuals identified as current smokers reported nondaily smoking patterns, with up to half categorized as occasional rather than daily smokers. To account for cumulative tobacco exposure, we also included smoking pack-years in the model.


**Extended acknowledgement**

Molecular data for the Trans-Omics in Precision Medicine (TOPMed) program was supported by the National Heart, Lung and Blood Institute (NHLBI). See the TOPMed Omics Support Table below for study specific omics support information.

| TOPMed accession number | TOPMed Project | Parent Study Name TOPMed | TOPMed Phase | Omics Center | Omics Support |
|---|---|---|---|---|---|
| phs001211 | VTE | ARIC | 2 | Baylor | 3U54HG003 273-12S2 / HHSN268201 500015C |
| phs001368 | CHS | CHS | 3 | Baylor | HHSN268201 600033I |
| phs000951 | COPD | COPDGene | 1 | NWGC | 3R01HL0898 56-08S1 |
| phs000951 | COPD | COPDGene | 2 | Broad | HHSN268201 500014C |
| phs000951 | COPD | COPDGene | 2.5 | Broad | HHSN268201 500014C |
| phs000974 | FHS | FHS | 1 | Broad | 3U54HG003 067-12S2 |
| phs001395 | HCHS/SOL | HCHS/SOL | 3 | Baylor | HHSN268201 600033I |
| phs000964 | JHS | JHS | 1 | NWGC | HHSN268201 100037C |
| phs001416 | MESA | MESA | 2 | Broad | 3U54HG003 067-13S1 |

**Cohort-specific acknowledgements**

**ARIC:** The Atherosclerosis Risk in Communities study has been funded in whole or in part with Federal funds from the National Heart, Lung, and Blood Institute, National Institutes of Health, Department of Health and Human Services (contract numbers HHSN268201700001I, HHSN268201700002I, HHSN268201700003I, HHSN268201700004I and HHSN268201700005I). The authors thank the staff and participants of the ARIC study for their important contributions. Whole genome sequencing (WGS) for the Trans-Omics in Precision Medicine (TOPMed) program was supported by the National Heart, Lung and Blood Institute (NHLBI). WGS for "NHLBI TOPMed: Atherosclerosis Risk in Communities (ARIC)" (phs001211) was performed at the Baylor College of Medicine Human Genome Sequencing Center (HHSN268201500015C and 3U54HG003273-12S2) and the Broad Institute for MIT and Harvard (3R01HL092577- 06S1). Centralized read mapping and genotype calling, along with variant quality metrics and filtering were provided by the TOPMed Informatics Research Center (3R01HL-117626-02S1). Phenotype harmonization, data management, sample-identity QC, and general study coordination, were provided by the TOPMed Data Coordinating Center (3R01HL- 120393- 02S1). We gratefully acknowledge the studies and participants who provided biological samples and data for TOPMed. The Genome Sequencing Program (GSP) was funded by the National Human Genome Research Institute (NHGRI), the National Heart, Lung, and Blood Institute (NHLBI), and the National Eye Institute (NEI). The GSP Coordinating Center (U24 HG008956) contributed to cross program scientific initiatives and provided logistical and general study coordination. The Centers for Common Disease Genomics (CCDG) program was supported by NHGRI and NHLBI, and whole genome sequencing was performed at the Baylor College of Medicine Human Genome Sequencing Center (UM1 HG008898).

**COPDGene:** This work was supported by NHLBI grants U01 HL089897 and U01 HL089856 and by NIH contract 75N92023D00011. The COPDGene study (NCT00608764) has also been supported by the COPD Foundation through contributions made to an Industry Advisory Committee that has included AstraZeneca, Bayer Pharmaceuticals, Boehringer-Ingelheim, Genentech, GlaxoSmithKline, Novartis, Pfizer, and Sunovion. A full listing of COPDGene investigators can be found at: http://www.copdgene.org/directory.

**CHS:** This Cardiovascular Health Study research was supported by NHLBI contracts HHSN268201200036C, HHSN268200800007C, HHSN268201800001C, N01HC55222, N01HC85079, N01HC85080, N01HC85081, N01HC85082, N01HC85083, N01HC85086, 75N92021D00006; and NHLBI grants U01HL080295, R01HL087652, R01HL103612, R01HL105756, R01HL120393, U01HL130114, and R01HL172803 with additional contribution from the National Institute of Neurological Disorders and Stroke (NINDS). Additional support was provided through R01AG023629 from the National Institute on Aging (NIA). A full list of principal CHS investigators and institutions can be found at CHS-NHLBI.org.

**HCHS/SOL:** The Hispanic Community Health Study/Study of Latinos is a collaborative study supported by contracts from the National Heart, Lung, and Blood Institute (NHLBI) to the

University of North Carolina (HHSN268201300001I / N01-HC-65233), University of Miami (HHSN268201300004I / N01-HC-65234), Albert Einstein College of Medicine (HHSN268201300002I / N01-HC-65235), University of Illinois at Chicago Revised 01/19/2024 Page 2 of 4 (HHSN268201300003I / N01- HC-65236 Northwestern Univ), and San Diego State University (HHSN268201300005I / N01-HC-65237). The following Institutes/Centers/Offices have contributed to the HCHS/SOL through a transfer of funds to the NHLBI: National Institute on Minority Health and Health Disparities ,National Institute on Deafness and Other Communication Disorders, National Institute of Dental and Craniofacial Research, National Institute of Diabetes and Digestive and Kidney Diseases, National Institute of Neurological Disorders and Stroke, NIH Institution-Office of Dietary Supplements.

**FHS:** The Framingham Heart Study (FHS) acknowledges the support of contracts NO1-HC-25195, HHSN268201500001I and 75N92019D00031 from the National Heart, Lung and Blood Institute and grant supplement R01 HL092577-06S1 for this research. We also acknowledge the dedication of the FHS study participants without whom this research would not be possible. Dr. Vasan is supported in part by the Evans Medical Foundation and the Jay and Louis Coffman Endowment from the Department of Medicine, Boston University School of Medicine.

**JHS:** The Jackson Heart Study (JHS) is supported and conducted in collaboration with Jackson State University (HHSN268201800013I), Tougaloo College (HHSN268201800014I), the Mississippi State Department of Health (HHSN268201800015I) and the University of Mississippi Medical Center (HHSN268201800010I, HHSN268201800011I, and HHSN268201800012I) contracts from the National Heart, Lung, and Blood Institute (NHLBI) and the National Institute on Minority Health and Health Disparities (NIMHD). The authors also wish to thank the staff and participants of the JHS.

**MESA:** Molecular data for the Trans-Omics in Precision Medicine (TOPMed) program was supported by the National Heart, Lung and Blood Institute (NHLBI). See the TOPMed Omics Support Table below for study specific omics support information. Core support including centralized genomic read mapping and genotype calling, along with variant quality metrics and filtering were provided by the TOPMed Informatics Research Center (3R01HL-117626-02S1; contract HHSN268201800002I). Core support including phenotype harmonization, data management, sample-identity QC, and general program coordination were provided by the TOPMed Data Coordinating Center (R01HL-120393; U01HL-120393; contract HHSN268201800001I). We gratefully acknowledge the studies and participants who provided biological samples and data for TOPMed.The MESA projects are conducted and supported by the National Heart, Lung, and Blood Institute (NHLBI) in collaboration with MESA investigators. Support for MESA is provided by contracts 75N92025D00022, 75N92020D00001, HHSN268201500003I, N01-HC-95159, 75N92025D00026, 75N92020D00005, N01-HC-95160, 75N92020D00002, N01-HC-95161, 75N92025D00024,

75N92020D00003, N01-HC-95162, 75N92025D00027, 75N92020D00006, N01-HC-95163, 75N92025D00025, 75N92020D00004, N01-HC-95164, 75N92025D00028, 75N92020D00007, N01-HC-95165, N01-HC-95166, N01-HC-95167, N01-HC-95168, N01-HC-95169, UL1-TR-000040, UL1-TR-001079, UL1-TR-001420, UL1TR001881, DK063491, and R01HL105756. The authors thank the MESA participants and the MESA investigators and staff for their valuable contributions.

**Supplementary Tables**

Supplementary Table 1 Descriptive statistics of the additional TOPMed population-based cohorts at baseline for cross-sectional analyses, including data from five cohort studies: ARIC, CHS, FHS, HCHS/SOL, and JHS.

| **Additional TOPMed population-based cohorts** | | **without mLOY** | **with mLOY** | **p** |
|---|---|---|---|---|
| sample size | total (%) | 4526 (85) | 819 (15) | |
| Cell Fractions (CF, %) | Median (IQR) | 0.0 (0.0 to 0.0) | 14.0 (8.9 to 24.0) | <0.001 |
| Age at blood draw (years) | Median (IQR) | 58.0 (49.0 to 67.0) | 71.4 (65.0 to 77.0) | <0.001 |
| Cohort | ARIC | 1145 (25.3) | 150 (18.3) | <0.001 |
| | CHS | 691 (15.3) | 402 (49.1) | |
| | FHS | 548 (12.1) | 139 (17.0) | |
| | HCHS_SOL | 1288 (28.5) | 69 (8.4) | |
| | JHS | 854 (18.9) | 59 (7.2) | |
| Self-reported race/ethnicity | Black or African American | 1022 (22.6) | 97 (11.8) | <0.001 |
| | Hispanics/Latinos | 1288 (28.5) | 69 (8.4) | |
| | Native American | 1 (0.0) | 0 (0.0) | |
| | Others | 5 (0.1) | 1 (0.1) | |
| | White | 2210 (48.8) | 652 (79.6) | |
| Smoking status | current smoker | 898 (19.9) | 157 (19.3) | <0.001 |
| | former smoker | 1853 (41.1) | 463 (56.8) | |
| | never smoker | 1758 (39.0) | 195 (23.9) | |
| Smoking pack-years | Median (IQR) | 4.6 (0.0 to 27.0) | 22.5 (0.0 to 48.0) | <0.001 |

| | | | | |
|---|---|---|---|---|
| Height (cm) | Median (IQR) | 175.0 (169.5 to 179.0) | 173.0 (168.9 to 177.3) | <0.001 |
| GOLD grade | 0 | 2920 (64.7) | 327 (40.1) | <0.001 |
| | -1 | 499 (11.1) | 68 (8.3) | |
| | 1 | 508 (11.3) | 189 (23.2) | |
| | 2 | 494 (10.9) | 168 (20.6) | |
| | 3 | 76 (1.7) | 50 (6.1) | |
| | 4 | 16 (0.4) | 13 (1.6) | |
| pre FEV1 | Median (IQR) | 3.1 (2.6 to 3.6) | 2.5 (2.0 to 3.0) | <0.001 |
| pre FVC | Median (IQR) | 4.1 (3.5 to 4.7) | 3.7 (3.1 to 4.2) | <0.001 |
| pre FEV1/FVC | Median (IQR) | 76.3 (70.2 to 81.3) | 69.6 (62.9 to 75.2) | <0.001 |
| FEV1 percent predicted - HANK | Median (IQR) | 0.9 (0.8 to 1.0) | 0.9 (0.7 to 1.0) | <0.001 |

Supplementary Table 2 Descriptive statistics of the additional TOPMed population-based cohorts at baseline for longitudinal and prospective analyses, including data from five cohort studies: ARIC, CHS, FHS, and MESA.

| **Additional TOPMed population-based cohorts** | | **without mLOY** | **with mLOY** | ***p*** |
|---|---|---|---|---|
| sample size | total (%) | 3080 (84) | 606 (16) | |
| Cell Fractions (CF) | Median (IQR) | 0.0 (0.0 to 0.0) | 14.3 (9.0 to 26.0) | <0.001 |
| Age at blood draw (years) | Median (IQR) | 60.6 (53.0 to 68.0) | 72.0 (65.0 to 77.0) | <0.001 |
| Follow-up time (years) | Median (IQR) | 5.0 (0.5 to 15.0) | 0.6 (0.0 to 5.0) | <0.001 |

| | | | | |
|---|---|---|---|---|
| Cohort | ARIC | 834 (27.1) | 121 (20.0) | <0.001 |
| | CHS | 332 (10.8) | 213 (35.1) | |
| | FHS | 211 (6.9) | 85 (14.0) | |
| | MESA | 1703 (55.3) | 187 (30.9) | |
| Self-reported race/ethnicity | Asian | 267 (8.7) | 21 (3.5) | <0.001 |
| | Black or African American | 521 (16.9) | 66 (10.9) | |
| | Hispanics/Latinos | 361 (11.7) | 34 (5.6) | |
| | Native American | 1 (0.0) | 0 (0.0) | |
| | Others | 2 (0.1) | 1 (0.2) | |
| | White | 1928 (62.6) | 484 (79.9) | |
| Smoking status | current smoker | 413 (13.5) | 103 (17.1) | <0.001 |
| | former smoker | 1731 (56.4) | 379 (62.9) | |
| | never smoker | 926 (30.2) | 121 (20.1) | |
| Smoking pack-years | Median (IQR) | 7.5 (0.0 to 30.0) | 23.5 (0.1 to 51.4) | <0.001 |
| Height (cm) | Median (IQR) | 173.5 (168.4 to 178.5) | 172.9 (168.0 to 177.0) | 0.013 |
| GOLD grade | Normal spirometry | 1483 (48.3) | 203 (33.8) | <0.001 |
| | PRISm | 468 (15.2) | 56 (9.3) | |
| | 1 | 461 (15.0) | 155 (25.8) | |
| | 2 | 553 (18.0) | 134 (22.3) | |
| | 3 | 88 (2.9) | 43 (7.2) | |

| | | | | |
|---|---|---|---|---|
| | 4 | 17 (0.6) | 10 (1.7) | |
| pre FEV1/FVC | Median (IQR) | 73.4 (67.7 to 78.1) | 69.9 (63.3 to 74.9) | <0.001 |
| FEV1 percent predicted - HANK | Median (IQR) | 0.9 (0.7 to 1.0) | 0.9 (0.7 to 1.0) | 0.171 |

Supplementary Table 3 Summary statistics of the cross-sectional association between mLOY and lung function across individual TOPMed population-based cohorts, adjusted for genetic ancestry principal components, age, study, age², smoking status, pack-years, height, and batch principal components. Asterisk (*) denotes a statistically significant association. Note that COPD, moderate COPD, severe COPD and PRISm are binary traits.

| **Cohort** | **Traits** | **Beta/OR [95%CI]** | ***p*** |
|---|---|---|---|
| ARIC | FEV1 percent predicted - Hank | -0.056 [-0.218, 0.106] | 0.5 |
| | GOLD grade (0 to 4) | 0.135 [-0.032, 0.302] | 0.113 |
| | pre FEV1 | -0.053 [-0.191, 0.085] | 0.453 |
| | pre FEV1/FVC | -0.188 [-0.350, -0.027] | 0.023* |
| | pre FVC | 0.045 [-0.092, 0.183] | 0.521 |
| | COPD | 1.566 [1.052, 2.331] | 0.027* |
| | moderate-to-severe COPD | 1.297 [0.764, 2.203] | 0.336 |
| | PRISm | 1.073 [0.459, 2.507] | 0.872 |
| | severe COPD | 1.298 [0.403, 4.188] | 0.662 |
| CHS | FEV1 percent predicted - Hank | -0.045 [-0.173, 0.082] | 0.489 |
| | GOLD grade (0 to 4) | 0.121 [-0.013, 0.256] | 0.077 |
| | pre FEV1 | -0.065 [-0.185, 0.055] | 0.288 |
| | pre FEV1/FVC | -0.157 [-0.285, -0.028] | 0.017* |
| | pre FVC | 0.022 [-0.094, 0.139] | 0.709 |
| | COPD | 1.255 [0.917, 1.719] | 0.157 |

| | | | |
|---|---|---|---|
| | moderate-to-severe COPD | 1.153 [0.795, 1.671] | 0.453 |
| | PRISm | 1.081 [0.622, 1.879] | 0.782 |
| | severe COPD | 1.746 [0.919, 3.316] | 0.089 |
| FHS | FEV1 percent predicted - Hank | -0.055 [-0.245, 0.134] | 0.567 |
| | GOLD grade (0 to 4) | 0.155 [-0.033, 0.343] | 0.106 |
| | pre FEV1 | -0.049 [-0.200, 0.103] | 0.528 |
| | pre FEV1/FVC | -0.201 [-0.384, -0.018] | 0.032* |
| | pre FVC | 0.045 [-0.106, 0.196] | 0.559 |
| | COPD | 1.243 [0.794, 1.947] | 0.341 |
| | moderate-to-severe COPD | 1.942 [0.972, 3.882] | 0.06 |
| | PRISm | 0.866 [0.284, 2.639] | 0.801 |
| HCHS/SOL | FEV1 percent predicted - Hank | -0.395 [-0.645, -0.144] | 0.002* |
| | GOLD grade (0 to 4) | 0.418 [0.160, 0.676] | 0.002* |
| | pre FEV1 | -0.228 [-0.407, -0.049] | 0.013* |
| | pre FEV1/FVC | -0.349 [-0.576, -0.121] | 0.003* |
| | pre FVC | -0.121 [-0.309, 0.067] | 0.208 |
| | COPD | 1.754 [0.947, 3.251] | 0.074 |
| | moderate-to-severe COPD | 2.386 [1.178, 4.832] | 0.016* |
| | PRISm | 1.039 [0.439, 2.459] | 0.931 |
| JHS | FEV1 percent predicted - Hank | -0.392 [-0.665, -0.119] | 0.005* |
| | GOLD grade (0 to 4) | 0.479 [0.165, 0.792] | 0.003* |
| | pre FEV1 | -0.262 [-0.473, -0.051] | 0.015* |
| | pre FEV1/FVC | -0.278 [-0.546, -0.010] | 0.042* |
| | pre FVC | -0.191 [-0.411, 0.028] | 0.087 |
| | COPD | 1.753 [0.778, 3.950] | 0.176 |

| | | | |
|---|---|---|---|
| | moderate-to-severe COPD | 2.415 [1.034, 5.645] | 0.042* |
| | PRISm | 1.667 [0.798, 3.481] | 0.174 |

| **Cohort** | **Traits** | **Beta/OR [95%CI]** | ***p*** |
|---|---|---|---|
| ARIC | FEV1 percent predicted - Hank | -0.056 [-0.218, 0.106] | 0.500 |
| | GOLD grade (0 to 4) | 0.135 [-0.032, 0.302] | 0.113 |
| | pre FEV1 | -0.053 [-0.191, 0.085] | 0.453 |
| | pre FEV1/FVC | -0.188 [-0.350, -0.027] | 0.023* |
| | pre FVC | 0.045 [-0.092, 0.183] | 0.521 |
| | COPD | 1.566 [1.052, 2.331] | 0.027* |
| | moderate-to-severe COPD | 1.297 [0.764, 2.203] | 0.336 |
| | PRISm | 1.073 [0.459, 2.507] | 0.872 |
| | severe COPD | 1.298 [0.403, 4.188] | 0.662 |
| CHS | FEV1 percent predicted - Hank | -0.045 [-0.173, 0.082] | 0.489 |
| | GOLD grade (0 to 4) | 0.121 [-0.013, 0.256] | 0.077 |
| | pre FEV1 | -0.065 [-0.185, 0.055] | 0.288 |
| | pre FEV1/FVC | -0.157 [-0.285, -0.028] | 0.017* |
| | pre FVC | 0.022 [-0.094, 0.139] | 0.709 |
| | COPD | 1.255 [0.917, 1.719] | 0.157 |
| | moderate-to-severe COPD | 1.153 [0.795, 1.671] | 0.453 |
| | PRISm | 1.081 [0.622, 1.879] | 0.782 |
| | severe COPD | 1.746 [0.919, 3.316] | 0.089 |
| FHS | FEV1 percent predicted - Hank | -0.055 [-0.245, 0.134] | 0.567 |
| | GOLD grade (0 to 4) | 0.155 [-0.033, 0.343] | 0.106 |
| | pre FEV1 | -0.049 [-0.200, 0.103] | 0.528 |
| | pre FEV1/FVC | -0.201 [-0.384, -0.018] | 0.032* |
| | pre FVC | 0.045 [-0.106, 0.196] | 0.559 |
| | COPD | 1.243 [0.794, 1.947] | 0.341 |
| | moderate-to-severe COPD | 1.942 [0.972, 3.882] | 0.060 |
| | PRISm | 0.866 [0.284, 2.639] | 0.801 |
| HCHS/SOL | FEV1 percent predicted - Hank | -0.395 [-0.645, -0.144] | 0.002* |
| | GOLD grade (0 to 4) | 0.418 [0.160, 0.676] | 0.002* |
| | pre FEV1 | -0.228 [-0.407, -0.049] | 0.013* |
| | pre FEV1/FVC | -0.349 [-0.576, -0.121] | 0.003* |

| | | | |
|---|---|---|---|
| | pre FVC | -0.121 [-0.309, 0.067] | 0.208 |
| | COPD | 1.754 [0.947, 3.251] | 0.074 |
| | moderate-to-severe COPD | 2.386 [1.178, 4.832] | 0.016* |
| | PRISm | 1.039 [0.439, 2.459] | 0.931 |
| JHS | FEV1 percent predicted - Hank | -0.392 [-0.665, -0.119] | 0.005* |
| | GOLD grade (0 to 4) | 0.479 [0.165, 0.792] | 0.003* |
| | pre FEV1 | -0.262 [-0.473, -0.051] | 0.015* |
| | pre FEV1/FVC | -0.278 [-0.546, -0.010] | 0.042* |
| | pre FVC | -0.191 [-0.411, 0.028] | 0.087 |
| | COPD | 1.753 [0.778, 3.950] | 0.176 |
| | moderate-to-severe COPD | 2.415 [1.034, 5.645] | 0.042* |
| | PRISm | 1.667 [0.798, 3.481] | 0.174 |

Supplementary Table 4 Summary statistics of associations between mLOY and lung function measures and COPD in COPDGene propensity age-matched set, adjusted for genetic ancestry principal components, age, age$^2$, smoking status, pack-years, height, and batch principal components. For LAA950 and log-transformed LAA950, adjustments also include scanner type. An asterisk (*) indicates a significant association. Note that COPD, moderate COPD, severe COPD and PRISm are binary traits.

| **Traits** | **OR/Beta [95% CI]** | ***p*** |
|---|---|---|
| COPD | 1.350 [1.036, 1.758] | 0.026* |
| moderate-to-severe COPD | 1.310 [0.996, 1.722] | 0.054 |
| PRISm | 1.082 [0.652, 1.797] | 0.761 |
| severe COPD | 1.277 [0.925, 1.762] | 0.137 |
| Dunedin Pace of Aging Methylation 38 | 0.239 [0.106, 0.372] | 4.74e-04* |
| FEV1 percent predicted - GLI GLOBAL | -0.116 [-0.212, -0.020] | 0.018* |
| FEV1/FVC percent predicted - GLI GLOBAL | -0.159 [-0.255, -0.062] | 0.001* |
| GOLD grade (0 to 4) | 0.089 [-0.011, 0.188] | 0.082 |
| LAA950 | 0.179 [0.051, 0.308] | 0.006* |

| | | |
|---|---|---|
| log-transformed LAA950 | 0.129 [0.005, 0.252] | 0.042* |
| post FEV1 percent predicted | -0.111 [-0.207, -0.014] | 0.025* |
| post FEV1/FVC | -0.156 [-0.252, -0.059] | 0.002* |
| pre FEV1/FVC | -0.140 [-0.238, -0.043] | 0.005* |

Supplementary Table 5 Summary statistics of associations between mLOY and lung function measures and COPD in additional TOPMed population-based cohorts using matched by age and study in the propensity score-matched set, adjusted for genetic ancestry principal components, age, age², smoking status, pack-years, height, and batch principal components. An asterisk (*) indicates a significant association. Note that COPD, moderate COPD, severe COPD and PRISm are binary traits.

| **Traits** | **Beta/OR [95% CI]** | ***p*** |
|---|---|---|
| FEV1 percent predicted - Hank | -0.056 [-0.148, 0.037] | 0.242 |
| GOLD grade (0 to 4) | 0.106 [0.006, 0.205] | 0.037* |
| pre FEV1 | -0.056 [-0.137, 0.025] | 0.178 |
| pre FEV1/FVC | -0.127 [-0.220, -0.033] | 0.008* |
| pre FVC | 0.016 [-0.062, 0.094] | 0.694 |
| COPD | 1.247 [0.983, 1.581] | 0.069 |
| moderate-to-severe COPD | 1.238 [0.923, 1.660] | 0.154 |
| PRISm | 1.033 [0.698, 1.529] | 0.872 |
| severe COPD | 1.647 [0.923, 2.938] | 0.091 |

Supplementary Table 6 Summary statistics of cross-sectional smoking status stratification association between mLOY and lung function in COPDGene, adjusted for genetic ancestry principal components, age, age², smoking status, pack-years, height, and batch principal components. For LAA950 and log-transformed LAA950, adjustments also include scanner type. An asterisk (*) indicates a significant association. Note that COPD, moderate COPD, severe COPD and PRISm are binary traits.

| **Smoking status** | **Traits** | **OR/Beta [95% CI]** | ***p*** |
|---|---|---|---|
| Former smokers | COPD | 1.445 [1.101, 1.898] | 0.008* |

| | | | |
|---|---|---|---|
| | moderate-to-severe COPD | 1.439 [1.086, 1.907] | 0.011* |
| | PRISm | 0.879 [0.503, 1.537] | 0.651 |
| | severe COPD | 1.427 [1.034, 1.969] | 0.031* |
| | Dunedin Pace of Aging Methylation 38 | 0.203 [0.052, 0.354] | 0.009* |
| | FEV1 percent predicted - GLI GLOBAL | -0.165 [-0.263, -0.068] | 9.18e-04* |
| | FEV1/FVC percent predicted - GLI GLOBAL | -0.211 [-0.309, -0.113] | 2.67e-05* |
| | GOLD grade (0 to 4) | 0.148 [0.047, 0.250] | 0.004* |
| | LAA950 | 0.219 [0.088, 0.350] | 0.001* |
| | log-transformed LAA950 | 0.144 [0.018, 0.271] | 0.026* |
| | post FEV1 percent predicted | -0.161 [-0.260, -0.062] | 0.001* |
| | post FEV1/FVC | -0.208 [-0.305, -0.110] | 2.98e-05* |
| | pre FEV1/FVC | -0.186 [-0.284, -0.088] | 2.04e-04* |
| Current smokers | COPD | 0.957 [0.682, 1.342] | 0.798 |
| | moderate-to-severe COPD | 0.826 [0.574, 1.189] | 0.304 |
| | PRISm | 0.802 [0.453, 1.418] | 0.448 |
| | severe COPD | 0.543 [0.314, 0.938] | 0.029* |
| | Dunedin Pace of Aging Methylation 38 | 0.168 [-0.039, 0.375] | 0.112 |
| | FEV1 percent predicted - GLI GLOBAL | 0.152 [0.021, 0.284] | 0.023* |
| | FEV1/FVC percent predicted - GLI GLOBAL | 0.077 [-0.047, 0.202] | 0.224 |
| | GOLD grade (0 to 4) | -0.123 [-0.254, 0.008] | 0.067 |
| | LAA950 | 0.027 [-0.149, 0.204] | 0.761 |
| | log-transformed LAA950 | 0.091 [-0.077, 0.258] | 0.289 |
| | post FEV1 percent predicted | 0.151 [0.021, 0.280] | 0.022* |
| | post FEV1/FVC | 0.075 [-0.047, 0.197] | 0.229 |

| | pre FEV1/FVC | 0.055 [-0.069, 0.178] | 0.387 |
|---|---|---|---|

Supplementary Table 7 Summary statistics of cross-sectional smoking status stratification association between mLOY and lung function in COPDGene propensity age-matched set**,** adjusted for genetic ancestry principal components, age, age², smoking status, pack-years, height, and batch principal components. For LAA950 and log-transformed LAA950, adjustments also include scanner type. An asterisk (*) indicates a significant association. Note that COPD, moderate COPD, severe COPD and PRISm are binary traits.

| **Smoking status** | **Traits** | **OR/Beta [95% CI]** | ***p*** |
|---|---|---|---|
| Former smokers | COPD | 1.717 [1.213, 2.430] | 0.002* |
| | moderate-to-severe COPD | 1.730 [1.210, 2.474] | 0.003* |
| | PRISm | 1.219 [0.602, 2.469] | 0.583 |
| | severe COPD | 1.678 [1.117, 2.521] | 0.013* |
| | Dunedin Pace of Aging Methylation 38 | 0.306 [0.125, 0.486] | 9.64e-04* |
| | FEV1 percent predicted - GLI GLOBAL | -0.202 [-0.314, -0.090] | 4.35e-04* |
| | FEV1/FVC percent predicted - GLI GLOBAL | -0.234 [-0.350, -0.119] | 7.23e-05* |
| | GOLD grade (0 to 4) | 0.169 [0.052, 0.286] | 0.005* |
| | LAA950 | 0.196 [0.040, 0.352] | 0.014* |
| | log-transformed LAA950 | 0.098 [-0.055, 0.251] | 0.209 |
| | post FEV1 percent predicted | -0.200 [-0.313, -0.086] | 5.90e-04* |
| | post FEV1/FVC | -0.234 [-0.349, -0.118] | 7.53e-05* |
| | pre FEV1/FVC | -0.199 [-0.315, -0.082] | 8.61e-04* |
| Current smokers | COPD | 0.854 [0.526, 1.388] | 0.525 |
| | moderate-to-severe COPD | 0.734 [0.435, 1.239] | 0.246 |
| | PRISm | 1.336 [0.515, 3.461] | 0.551 |
| | severe COPD | 0.555 [0.271, 1.139] | 0.108 |
| | Dunedin Pace of Aging Methylation 38 | 0.141 [-0.138, 0.420] | 0.324 |
| | FEV1 percent predicted - GLI GLOBAL | 0.110 [-0.069, 0.288] | 0.229 |

| | | |
|---|---|---|
| FEV1/FVC percent predicted - GLI GLOBAL | 0.031 [-0.140, 0.203] | 0.720 |
| GOLD grade (0 to 4) | -0.112 [-0.293, 0.069] | 0.227 |
| LAA950 | 0.083 [-0.170, 0.336] | 0.521 |
| log-transformed LAA950 | 0.146 [-0.100, 0.393] | 0.246 |
| post FEV1 percent predicted | 0.100 [-0.077, 0.278] | 0.267 |
| post FEV1/FVC | 0.031 [-0.138, 0.201] | 0.717 |
| pre FEV1/FVC | 0.016 [-0.159, 0.191] | 0.857 |

Supplementary Table 8 Summary statistics of cross-sectional smoking status stratification association between mLOY and lung function in additional TOPMed population-based cohorts, adjusted for genetic ancestry principal components, age, age², study, smoking status, pack-years, height, and batch principal components. An asterisk (*) indicates a significant association. Note that COPD, moderate COPD, severe COPD and PRISm are binary traits.

| **Smoking status** | **Traits** | **OR/Beta [95% CI]** | ***p*** |
|---|---|---|---|
| Current smokers | FEV1 percent predicted - Hank | -0.353 [-0.530, -0.176] | 1.03e-04* |
| | GOLD grade (0 to 4) | 0.371 [0.196, 0.547] | 3.69e-05* |
| | pre FEV1/FVC | -0.322 [-0.478, -0.166] | 5.86e-05* |
| | COPD | 2.154 [1.349, 3.437] | 0.001* |
| | moderate-to-severe COPD | 2.090 [1.238, 3.531] | 0.006* |
| | PRISm | 1.476 [0.698, 3.122] | 0.309 |
| | severe COPD | 2.313 [0.937, 5.706] | 0.069 |
| Former smokers | FEV1 percent predicted - Hank | -0.117 [-0.225, -0.009] | 0.034* |
| | GOLD grade (0 to 4) | 0.165 [0.055, 0.276] | 0.003* |
| | pre FEV1/FVC | -0.186 [-0.287, -0.084] | 3.40e-04* |
| | COPD | 1.285 [0.988, 1.670] | 0.062 |
| | moderate-to-severe COPD | 1.448 [1.053, 1.991] | 0.023* |
| | PRISm | 1.059 [0.683, 1.642] | 0.797 |
| | severe COPD | 2.432 [1.333, 4.438] | 0.004* |

| | | | |
|---|---|---|---|
| Never smokers | FEV1 percent predicted - Hank | 0.175 [0.016, 0.333] | 0.031* |
| | GOLD grade (0 to 4) | -0.001 [-0.163, 0.161] | 0.99 |
| | pre FEV1/FVC | -0.050 [-0.191, 0.090] | 0.483 |
| | COPD | 1.183 [0.804, 1.742] | 0.393 |
| | moderate-to-severe COPD | 0.892 [0.500, 1.590] | 0.698 |
| | PRISm | 0.725 [0.361, 1.457] | 0.366 |

Supplementary Table 9 Summary statistics of cross-sectional smoking status stratification association between mLOY and lung function in additional TOPMed population-based cohorts using matched by age and study in the propensity score-matched set, adjusted for genetic ancestry principal components, age, age², study, smoking status, pack-years, height, and batch principal components. An asterisk (*) indicates a significant association. Note that COPD, moderate COPD, severe COPD and PRISm are binary traits.

| **Smoking status** | **Traits** | **OR/Beta [95% CI]** | ***p*** |
|---|---|---|---|
| Current smokers | FEV1 percent predicted - Hank | -0.290 [-0.569, -0.011] | 0.043* |
| | GOLD grade (0 to 4) | 0.344 [0.042, 0.645] | 0.027* |
| | pre FEV1/FVC | -0.288 [-0.560, -0.017] | 0.039* |
| | COPD | 1.901 [0.907, 3.985] | 0.089 |
| | moderate-to-severe COPD | 2.484 [1.031, 5.986] | 0.043* |
| | PRISm | 1.378 [0.269, 7.070] | 0.701 |
| | Severe COPD | 5.105 [0.283, 92.218] | 0.270 |
| Former smokers | FEV1 percent predicted - Hank | -0.101 [-0.229, 0.027] | 0.124 |
| | GOLD grade (0 to 4) | 0.077 [-0.060, 0.213] | 0.272 |
| | pre FEV1/FVC | -0.112 [-0.241, 0.017] | 0.09 |
| | COPD | 1.076 [0.785, 1.474] | 0.649 |
| | moderate-to-severe COPD | 1.137 [0.779, 1.662] | 0.505 |
| | PRISm | 1.078 [0.639, 1.819] | 0.778 |
| | severe COPD | 1.773 [0.827, 3.801] | 0.141 |

| | | | |
|---|---|---|---|
| Never smokers | FEV1 percent predicted - Hank | 0.170 [-0.014, 0.353] | 0.072 |
| | GOLD grade (0 to 4) | 0.011 [-0.190, 0.213] | 0.913 |
| | pre FEV1/FVC | -0.041 [-0.227, 0.145] | 0.664 |
| | COPD | 1.318 [0.825, 2.107] | 0.248 |
| | moderate-to-severe COPD | 0.975 [0.498, 1.910] | 0.942 |
| | PRISm | 0.658 [0.267, 1.620] | 0.363 |

Supplementary Table 10 Summary statistics of mLOY distribution across each age group categorisation in COPDGene for cross-sectional age-stratification analysis in Supplementary Table 11.

| | | **without mLOY** | **with mLOY** | **p** |
|---|---|---|---|---|
| | Age at blood draw (years) | **Sample size (%)** | | |
| | < 50 | 846 (19.5) | 9 (1.2) | <0.001 |
| | ≥ 50, < 60 | 1772 (40.7) | 100 (13.4) | |
| | ≥ 60, < 70 | 1225 (28.2) | 321 (42.9) | |
| | ≥ 70 | 506 (11.6) | 318 (42.5) | |
| < 50 | Median (IQR) | 47.6 (46.3 to 48.8) | 48.4 (47.1 to 48.7) | |
| smoking status | Current smoker | 712 (84.2) | 8 (88.9) | |
| | Former smoker | 134 (15.8) | 1 (11.1) | |
| ≥ 50, < 60 | Median (IQR) | 54.5 (52.2 to 57.3) | 56.6 (54.3 to 58.2) | |
| smoking status | Current smoker | 1299 (73.3) | 77 (77.0) | |
| | Former smoker | 473 (26.7) | 23 (23.0) | |
| ≥ 60, < 70 | Median (IQR) | 64.1 (62.0 to 66.7) | 65.5 (63.0 to 67.7) | |
| smoking status | Current smoker | 439 (35.8) | 125 (38.9) | |
| | Former smoker | 786 (64.2) | 196 (61.1) | |

|  |  |  |  |
| --- | --- | --- | --- |
| ≥ 70 | Median (IQR) | 73.1 (71.5 to 75.8) | 74.6 (72.1 to 77.3) |
| smoking status | Current smoker | 81 (16.0) | 41 (12.9) |
|  | Former smoker | 425 (84.0) | 277 (87.1) |

Supplementary Table 11 Summary statistics of age-stratification association between mLOY and lung function in COPDGene, adjusted for genetic ancestry principal components, age, age², smoking status, pack-years, height, and batch principal components. For LAA950 and log-transformed LAA950, adjustments also include scanner type. An asterisk (*) indicates a significant association. The summary statistics of mLOY distribution across each age group can be found in Supplementary Table 10. Note that COPD, moderate COPD, severe COPD and PRISm are binary traits.

| **Age group** | **Traits** | **OR/Beta [95% CI]** | ***p*** |
| --- | --- | --- | --- |
| ≥ 50, < 60 | COPD | 1.396 [0.876, 2.226] | 0.161 |
|  | Dunedin Pace of Aging Methylation 38 | 0.618 [0.356, 0.881] | 4.61e-06* |
|  | FEV1 percent predicted - GLI GLOBAL | -0.048 [-0.235, 0.139] | 0.616 |
|  | FEV1/FVC percent predicted - GLI GLOBAL | -0.023 [-0.205, 0.159] | 0.805 |
|  | GOLD grade (0 to 4) | 0.072 [-0.123, 0.267] | 0.469 |
|  | LAA950 | 0.177 [-0.071, 0.424] | 0.162 |
|  | log-transformed LAA950 | 0.192 [-0.047, 0.432] | 0.116 |
|  | post FEV1 percent predicted | -0.025 [-0.213, 0.162] | 0.791 |
|  | post FEV1/FVC | -0.020 [-0.201, 0.161] | 0.827 |
|  | pre FEV1/FVC | -0.076 [-0.262, 0.110] | 0.425 |
| ≥ 60, < 70 | COPD | 1.201 [0.879, 1.640] | 0.250 |
|  | Dunedin Pace of Aging Methylation 38 | 0.135 [-0.025, 0.295] | 0.099 |

| | | | |
|---|---|---|---|
| | FEV1 percent predicted - GLI GLOBAL | -0.013 [-0.131, 0.105] | 0.825 |
| | FEV1/FVC percent predicted - GLI GLOBAL | -0.114 [-0.233, 0.005] | 0.060 |
| | GOLD grade (0 to 4) | -0.005 [-0.129, 0.119] | 0.939 |
| | LAA950 | 0.152 [0.003, 0.302] | 0.046* |
| | log-transformed LAA950 | 0.210 [0.063, 0.356] | 0.005* |
| | post FEV1 percent predicted | -0.010 [-0.129, 0.110] | 0.875 |
| | post FEV1/FVC | -0.114 [-0.233, 0.004] | 0.059 |
| | pre FEV1/FVC | -0.101 [-0.221, 0.019] | 0.098 |
| ≥ 70 | COPD | 1.065 [0.631, 1.798] | 0.813 |
| | Dunedin Pace of Aging Methylation 38 | 5.35e-04 [-0.213, 0.214] | 0.996 |
| | FEV1 percent predicted - GLI GLOBAL | -0.116 [-0.240, 0.007] | 0.065 |
| | FEV1/FVC percent predicted - GLI GLOBAL | -0.128 [-0.257, 0.001] | 0.053 |
| | GOLD grade (0 to 4) | 0.094 [-0.032, 0.220] | 0.143 |
| | LAA950 | 0.143 [-0.043, 0.328] | 0.133 |
| | log-transformed LAA950 | -0.022 [-0.202, 0.158] | 0.812 |
| | post FEV1 percent predicted | -0.117 [-0.241, 0.007] | 0.066 |
| | post FEV1/FVC | -0.129 [-0.258, 7.86e-04] | 0.052 |
| | pre FEV1/FVC | -0.102 [-0.234, 0.029] | 0.128 |

Supplementary Table 12 Summary statistics of mLOY distribution across each age group categorisation in additional TOPMed population-based cohorts for cross-sectional age-stratification analysis in Supplementary Table 13.
* We did not summarize the group under 50 years old due to an insufficient number of individuals with mLOY.

| | | **without mLOY** | **with mLOY** | ***p*** |
|---|---|---|---|---|
| | Age at blood draw (years) | **sample size, %** | | |
| | <50* | 1219 (26.9) | 4 (0.5) | <0.001 |
| | ≥ 50, < 60 | 1278 (28.2) | 86 (10.5) | |
| | ≥ 60, < 70 | 1222 (27.0) | 260 (31.7) | |
| | ≥ 70 | 807 (17.8) | 469 (57.3) | |
| ≥ 50, < 60 | Median (IQR) | 55.0 (52.0 to 57.0) | 57.0 (54.1 to 58.0) | |
| Smoking status | current smoker | 309 (24.3) | 43 (50.0) | |
| | former smoker | 520 (41.0) | 29 (33.7) | |
| | never smoker | 440 (34.7) | 14 (16.3) | |
| ≥ 60, < 70 | Median (IQR) | 64.1 (62.0 to 67.0) | 66.0 (63.0 to 67.5) | |
| Smoking status | current smoker | 189 (15.5) | 65 (25.2) | |
| | former smoker | 646 (52.9) | 137 (53.1) | |
| | never smoker | 386 (31.6) | 56 (21.7) | |
| ≥ 70 | Median (IQR) | 74.0 (72.0 to 78.0) | 76.0 (73.0 to 80.0) | |
| Smoking status | current smoker | 57 (7.1) | 45 (9.6) | |
| | former smoker | 493 (61.3) | 297 (63.6) | |
| | never smoker | 254 (31.6) | 125 (26.8) | |

Supplementary Table 13 Summary statistics of age-stratification association between mLOY and lung function in additional TOPMed population-based cohorts, adjusted for genetic ancestry principal components, age, age$^2$, study, smoking status, pack-years, height, and batch principal components. An asterisk (*) indicates a significant association. The summary statistics of mLOY distribution across each age group can be found in Supplementary Table 12. Note that COPD, moderate COPD, severe COPD and PRISm are binary traits.

| Age at blood draw (years) groups | Traits | Beta/OR [95% CI] | *p* |
|---|---|---|---|
| ≥ 50, < 60 | COPD | 3.224 [1.919, 5.415] | 9.67e-06* |
| | FEV1 percent predicted - Hank | -0.379 [-0.588, -0.169] | 4.14e-04* |
| | GOLD grade (0 to 4) | 0.495 [0.271, 0.718] | 1.54e-05* |
| | pre FEV1/FVC | -0.298 [-0.501, -0.095] | 0.004* |
| ≥ 60, < 70 | COPD | 1.264 [0.913, 1.751] | 0.158 |
| | FEV1 percent predicted - Hank | -0.124 [-0.253, 0.004] | 0.059 |
| | GOLD grade (0 to 4) | 0.145 [0.014, 0.277] | 0.030* |
| | pre FEV1/FVC | -0.226 [-0.350, -0.103] | 3.48e-04* |
| ≥ 70 | COPD | 1.225 [0.923, 1.627] | 0.16 |
| | FEV1 percent predicted - Hank | -0.016 [-0.128, 0.096] | 0.781 |
| | GOLD grade (0 to 4) | 0.115 [-0.006, 0.236] | 0.063 |
| | pre FEV1/FVC | -0.121 [-0.236, -0.006] | 0.040* |

Supplementary Table 14 Summary statistics from sensitivity analysis of prospective associations between mLOY and lung function measures in participants with normal spirometry at the baseline in COPDGene, respectively, adjusted for genetic ancestry PCs, age, age$^2$, smoking status, pack-years, height, and batch effects. Both LAA950 and log-transformed LAA950 are additionally adjusted for scanner type. Significant associations are marked with an asterisk (*). Note that COPD, moderate COPD, severe COPD and PRISm are binary traits.

| Traits | OR/Beta [95% CI] | *p* |
|---|---|---|
| Dunedin Pace of Aging Methylation 38 | 0.330 [0.137, 0.523] | 8.25e-04* |
| FEV1 percent predicted - GLI GLOBAL | -0.181 [-0.361, 1.51e-04] | 0.050 |
| FEV1/FVC percent predicted - GLI GLOBAL | -0.171 [-0.366, 0.023] | 0.085 |
| GOLD grade (0 to 4) | 0.260 [0.055, 0.466] | 0.013* |
| LAA950 | -0.017 [-0.218, 0.184] | 0.867 |

| | | |
|---|---|---|
| log-transformed LAA950 | 0.018 [-0.175, 0.211] | 0.855 |
| post FEV1 percent predicted | -0.200 [-0.396, -0.005] | 0.045* |
| post FEV1/FVC | -0.172 [-0.362, 0.017] | 0.075 |
| pre FEV1/FVC | -0.147 [-0.340, 0.047] | 0.137 |
| COPD | 1.837 [1.100, 3.066] | 0.020* |
| moderate-to-severe COPD | 2.166 [0.902, 5.202] | 0.084 |
| PRISm | 1.525 [0.730, 3.186] | 0.261 |

Supplementary Table 15 Summary statistics from sensitivity analysis of prospective associations between mLOY and lung function measures in participants with normal spirometry at the baseline in additional TOPMed population-based cohorts, respectively, adjusted for genetic ancestry PCs, age, age$^2$, study, smoking status, pack-years, height, and batch effects. Significant associations are marked with an asterisk (*). Note that COPD, moderate COPD, severe COPD and PRISm are binary traits.

| **Traits** | **Beta [95% CI]** | ***p*** |
|---|---|---|
| FEV1 percent predicted - Hank | -0.024 [-0.225, 0.176] | 0.812 |
| GOLD grade (0 to 4) | -0.078 [-0.301, 0.146] | 0.497 |
| pre FEV1/FVC | -0.259 [-0.573, 0.056] | 0.108 |
| COPD | 0.813 [0.425, 1.558] | 0.534 |
| moderate-to-severe COPD | 0.713 [0.180, 2.814] | 0.629 |
| PRISm | 2.868 [1.089, 7.555] | 0.033* |

Supplementary Table 16 Summary statistics from the sensitivity analysis of cross-sectional data in COPDGene, where individuals with mLOY were classified as having a cell fraction ≥10%. Adjusted for genetic ancestry PCs, age, age$^2$, smoking status, pack-years, height, and batch effects. For LAA950 and log-transformed LAA950, adjustments also include scanner type. Significant associations are marked with an asterisk (*). Note that COPD, moderate COPD, severe COPD and PRISm are binary traits.

| **Traits** | **OR/Beta [95% CI]** | ***p*** |
|---|---|---|
| COPD | 1.239 [0.973, 1.577] | 0.082 |
| moderate-to-severe COPD | 1.173 [0.913, 1.506] | 0.212 |
| PRISm | 0.778 [0.481, 1.258] | 0.306 |

| | | |
|---|---|---|
| severe COPD | 1.209 [0.897, 1.629] | 0.213 |
| Dunedin Pace of Aging Methylation 38 | 0.211 [0.081, 0.341] | 0.002* |
| FEV1 percent predicted - GLI GLOBAL | -0.101 [-0.192, -0.011] | 0.028* |
| FEV1/FVC percent predicted - GLI GLOBAL | -0.165 [-0.252, -0.077] | 2.21e-04* |
| GOLD grade (0 to 4) | 0.076 [-0.014, 0.167] | 0.098 |
| LAA950 | 0.193 [0.077, 0.309] | 0.001* |
| log-transformed LAA950 | 0.129 [0.019, 0.238] | 0.021* |
| post FEV1 percent predicted | -0.088 [-0.177, 0.001] | 0.054 |
| post FEV1/FVC | -0.160 [-0.245, -0.075] | 2.36e-04* |
| pre FEV1/FVC | -0.144 [-0.230, -0.058] | 0.001* |

Supplementary Table 17 Summary statistics from the sensitivity analysis of cross-sectional data in additional TOPMed population-based cohorts, where individuals with mLOY were classified as having a cell fraction ≥10%. Adjusted for genetic ancestry PCs, age, age$^2$, study, smoking status, pack-years, height, and batch effects. Significant associations are marked with an asterisk (*). Note that COPD, moderate COPD, severe COPD and PRISm are binary traits.

| **Traits** | **Beta/OR [95% CI]** | **p** |
|---|---|---|
| FEV1 percent predicted - Hank | -0.126 [-0.218, -0.034] | 0.007* |
| GOLD grade (0 to 4) | 0.192 [0.102, 0.283] | 3.23e-05* |
| pre FEV1/FVC | -0.213 [-0.294, -0.132] | 2.50e-07* |
| COPD | 1.335 [1.065, 1.674] | 0.012* |
| moderate-to-severe COPD | 1.429 [1.085, 1.882] | 0.011* |
| PRISm | 1.078 [0.730, 1.590] | 0.706 |
| severe COPD | 2.091 [1.250, 3.497] | 0.005* |

Supplementary Table 18 Summary statistics from the sensitivity analysis of cross-sectional data in COPDGene, where aging-related markers, CHIP status and telomere length, were added to the primary model. Models were adjusted for genetic ancestry PCs, age, age$^2$, smoking status, pack-years, height, and batch effects. For LAA950 and log-transformed LAA950, adjustments also included scanner type. Significant associations are denoted with an asterisk (*). Note that COPD, moderate COPD, severe COPD and PRISm are binary traits.

| Additional confounder | Traits | OR/Beta [95% CI] | *p* |
|---|---|---|---|
| Telomere length | COPD | 1.185 [0.961, 1.460] | 0.112 |
| | moderate-to-severe COPD | 1.118 [0.899, 1.390] | 0.316 |
| | PRISm | 0.854 [0.577, 1.263] | 0.428 |
| | severe COPD | 1.059 [0.813, 1.379] | 0.672 |
| | Dunedin Pace of Aging Methylation 38 | 0.159 [0.048, 0.270] | 0.005* |
| | FEV1 percent predicted - GLI GLOBAL | -0.061 [-0.139, 0.017] | 0.127 |
| | FEV1/FVC percent predicted - GLI GLOBAL | -0.116 [-0.192, -0.040] | 0.003* |
| | GOLD grade (0 to 4) | 0.043 [-0.036, 0.121] | 0.287 |
| | LAA950 | 0.164 [0.062, 0.266] | 0.002* |
| | log-transformed LAA950 | 0.118 [0.023, 0.213] | 0.015* |
| | post FEV1 percent predicted | -0.050 [-0.127, 0.027] | 0.205 |
| | post FEV1/FVC | -0.112 [-0.186, -0.038] | 0.003* |
| | pre FEV1/FVC | -0.105 [-0.180, -0.031] | 0.006* |
| CHIP status | COPD | 1.259 [1.001, 1.584] | 0.049* |
| | moderate-to-severe COPD | 1.202 [0.947, 1.526] | 0.131 |
| | PRISm | 0.888 [0.575, 1.373] | 0.594 |
| | severe COPD | 1.171 [0.874, 1.568] | 0.290 |
| | Dunedin Pace of Aging Methylation 38 | 0.199 [0.079, 0.318] | 0.001* |
| | FEV1 percent predicted - GLI GLOBAL | -0.088 [-0.172, -0.004] | 0.040* |
| | FEV1/FVC percent predicted - GLI GLOBAL | -0.146 [-0.227, -0.065] | 3.84e-04* |
| | GOLD grade (0 to 4) | 0.074 [-0.010, 0.157] | 0.084 |
| | LAA950 | 0.161 [0.054, 0.269] | 0.003* |

| | | |
|---|---|---|
| log-transformed LAA950 | 0.113 [0.012, 0.215] | 0.028* |
| post FEV1 percent predicted | -0.078 [-0.160, 0.005] | 0.065 |
| post FEV1/FVC | -0.141 [-0.220, -0.063] | 4.43e-04* |
| pre FEV1/FVC | -0.144 [-0.223, -0.065] | 3.74e-04* |

**Supplementary Figures**

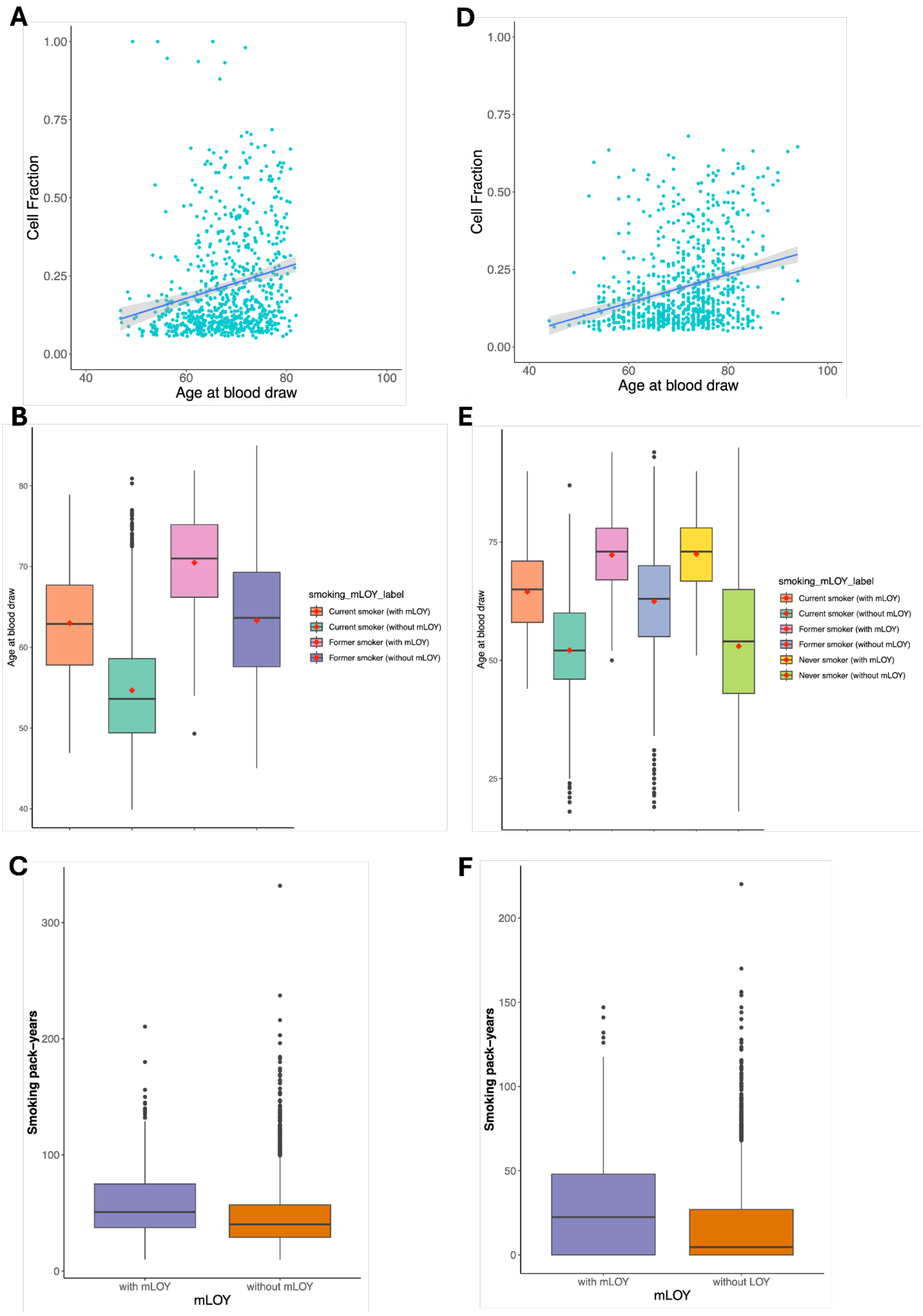


Supplementary Figure 1 A,D) Scatter plots showing the correlation between cell fraction estimates and age at blood draw in COPDGene and additional TOPMed population-based cohorts, respectively. B, E) Boxplots depicting the distribution of age at blood draw across mLOY and smoking status in COPDGene and additional TOPMed population-based cohorts, respectively. The red diamond shape indicates the mean of age at blood draw. C, F) Boxplots illustrating the distribution of smoking pack-years in individuals with and without mLOY in COPDGene and additional TOPMed population-based cohorts, respectively.

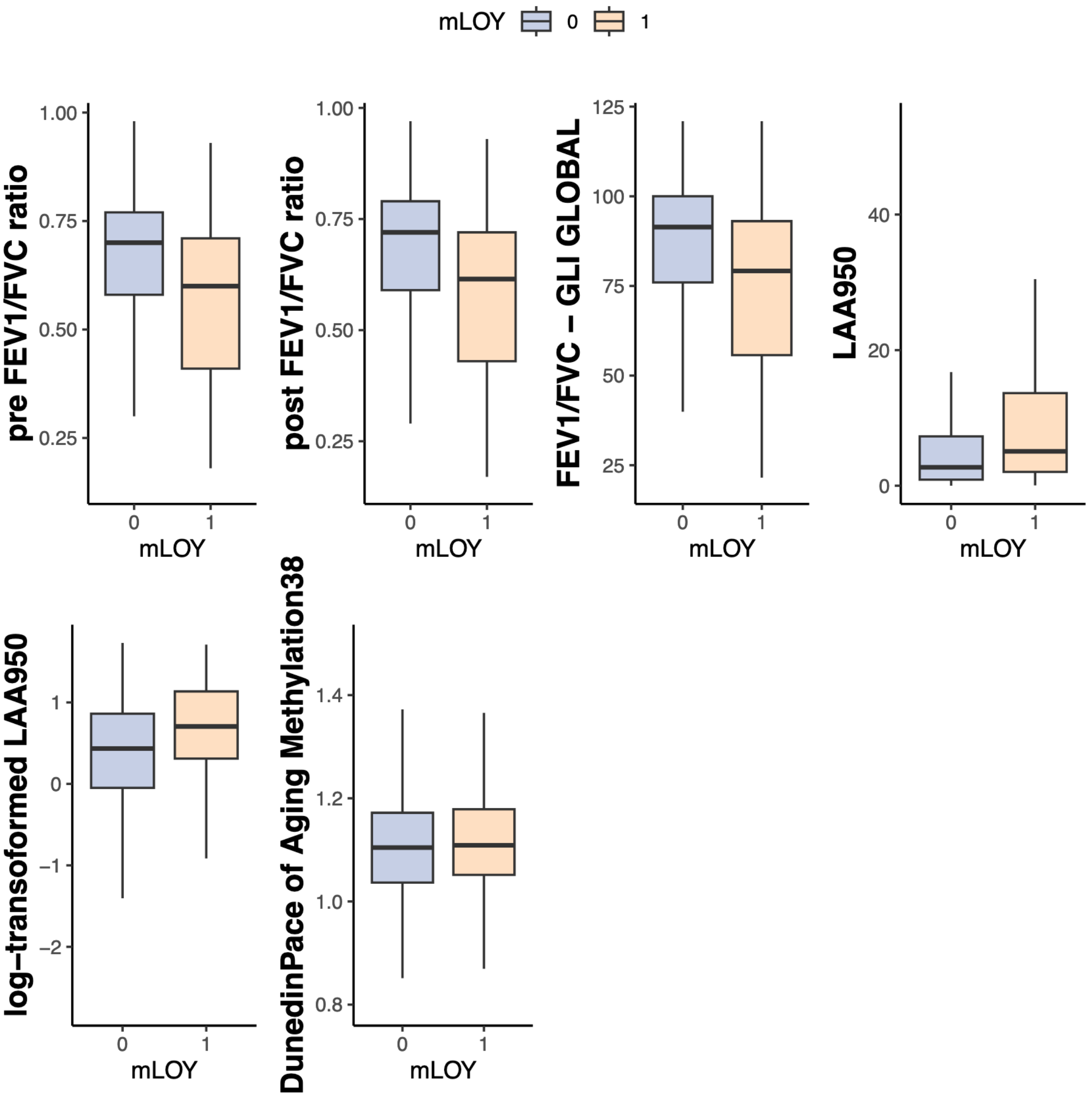


Supplementary Figure 2 Boxplots illustrating the distribution of traits significantly associated with mosaic loss of Y chromosome (mLOY) from the cross-sectional study are presented.Traits including pre-bronchodilator FEV1/FVC ratio, post-bronchodilator FEV1/FVC ratio, FEV1/FVC percent predicted – GLI GLOBAL, both LAA950 or log-transformed of emphysema indicator, and age acceleration. Each boxplot compares the distribution of these traits between individuals with mLOY (indicates as 1) and without mLOY (indicates as 0). Detailed statistical results and summaries can be found in main text Table 2.

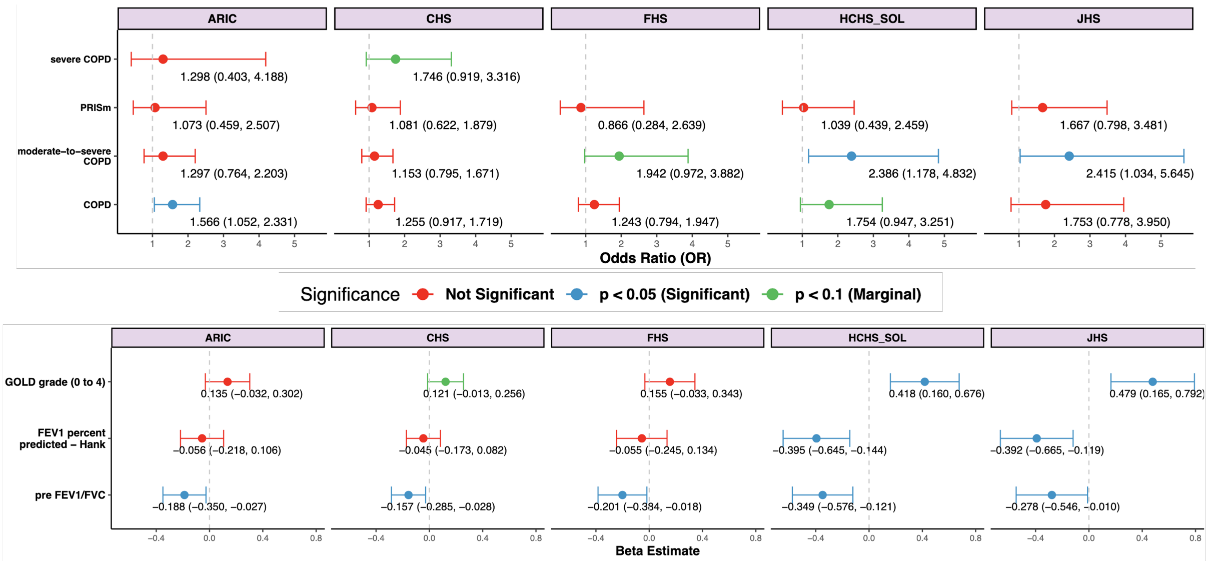


Supplementary Figure 3 Forest plots displaying beta coefficients for 95% confidence intervals (CI) for each single TOPMed cohort study in the cross-sectional analysis. These are shown for genetic ancestry principal components (PCs) adjustment, along with other confounders listed in the results section. Significance is color-coded. This analysis highlights that the association between mLOY and pre-bronchodilator FEV1/FVC, as reported in the main results and Figure 3, is not driven by any single cohort. Note that COPD, moderate COPD, severe COPD and PRISm are binary traits.

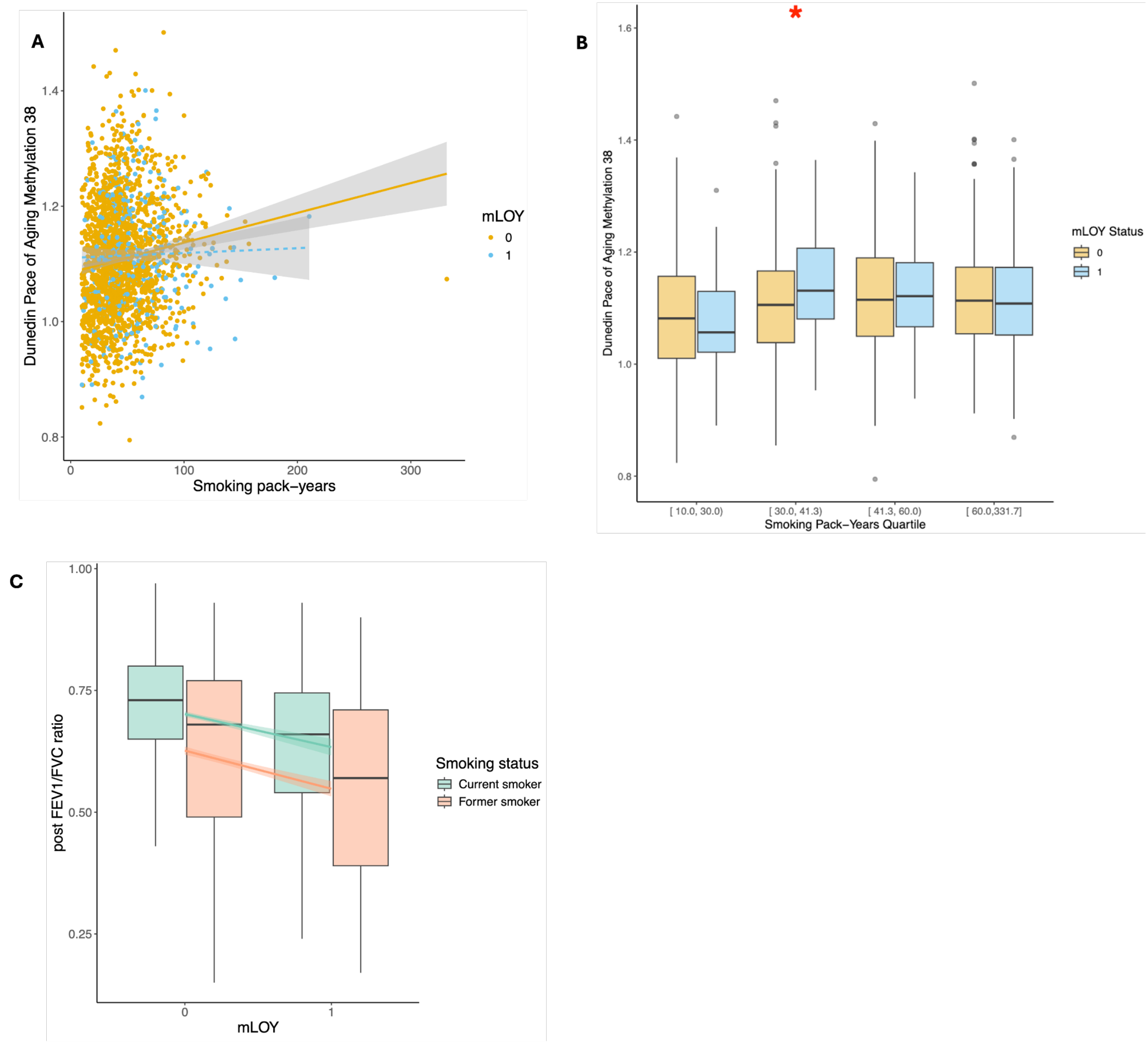


Supplementary Figure 4 A) Scatter plot showing the relationship between age acceleration (Y-axis) and smoking pack-years (X-axis), grouped by mLOY status (predictor). Each point represents an individual, color-coded by mLOY status; B) Boxplot illustrating smoking pack-years, divided into quartiles; C) Boxplot illustrating lung function (Y-axis) by mLOY status (X-axis), split by smoking status (interaction). The plot displays the distribution of lung function measures for both groups of individuals with mLOY and without mLOY, with separate boxes for current and former smokers, highlighting the combined effects of mLOY and smoking status on lung function. The mLOY status is indicated as 0 for individuals without mLOY and 1 for individuals with mLOY.

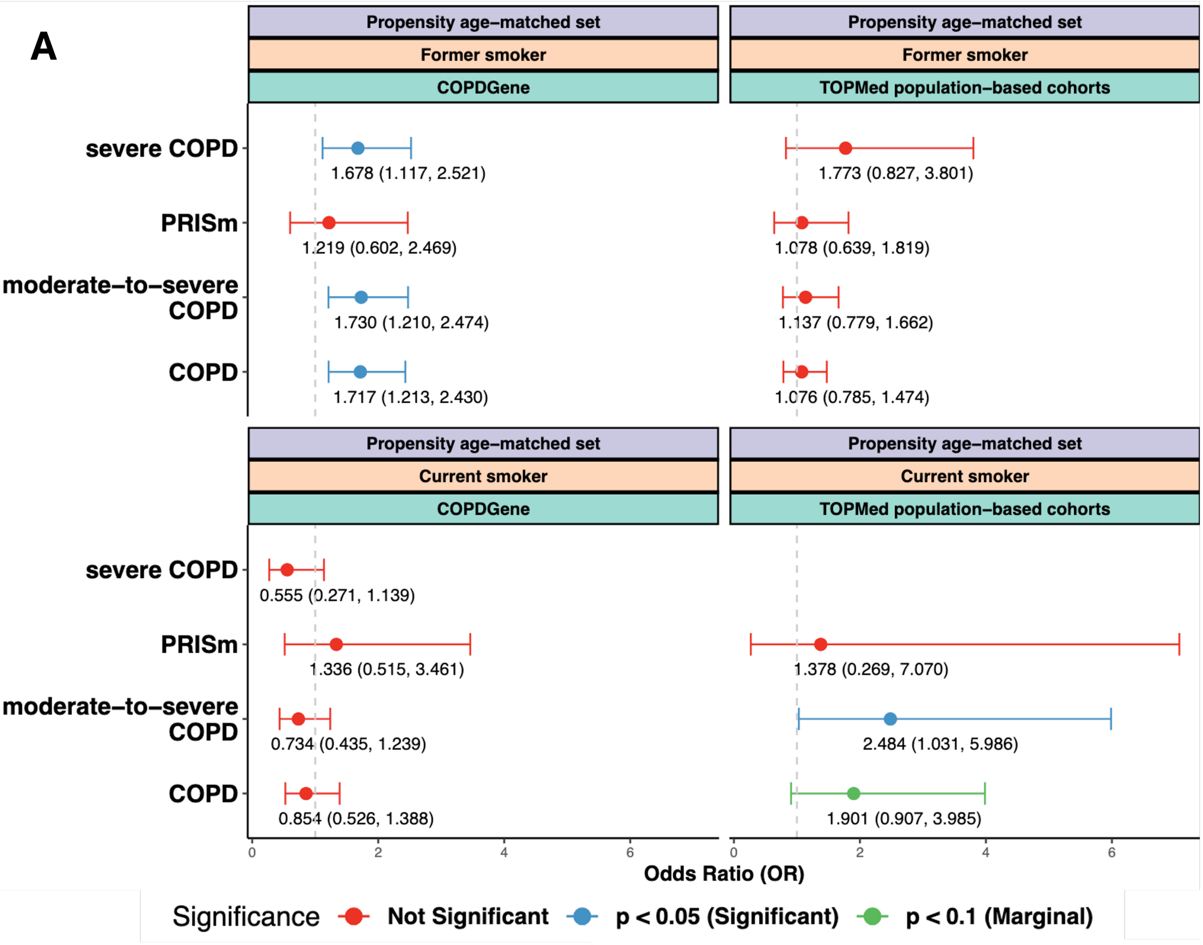

A
Propensity age–matched set
Former smoker
COPDGene
TOPMed population–based cohorts
Current smoker
severe COPD
PRISm
moderate–to–severe COPD
COPD
1.678 (1.117, 2.521)
1.219 (0.602, 2.469)
1.730 (1.210, 2.474)
1.717 (1.213, 2.430)
1.773 (0.827, 3.801)
1.078 (0.639, 1.819)
1.137 (0.779, 1.662)
1.076 (0.785, 1.474)
0.555 (0.271, 1.139)
1.336 (0.515, 3.461)
0.734 (0.435, 1.239)
0.854 (0.526, 1.388)
1.378 (0.269, 7.070)
2.484 (1.031, 5.986)
1.901 (0.907, 3.985)
0
2
4
6
Odds Ratio (OR)
Significance
Not Significant
p < 0.05 (Significant)
p < 0.1 (Marginal)


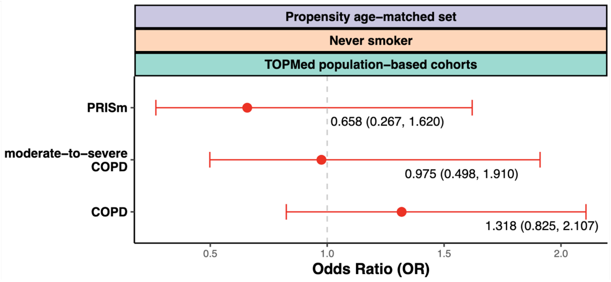

Propensity age–matched set
Never smoker
TOPMed population–based cohorts
PRISm
moderate–to–severe COPD
COPD
0.658 (0.267, 1.620)
0.975 (0.498, 1.910)
1.318 (0.825, 2.107)
0.5
1.0
1.5
2.0
Odds Ratio (OR)

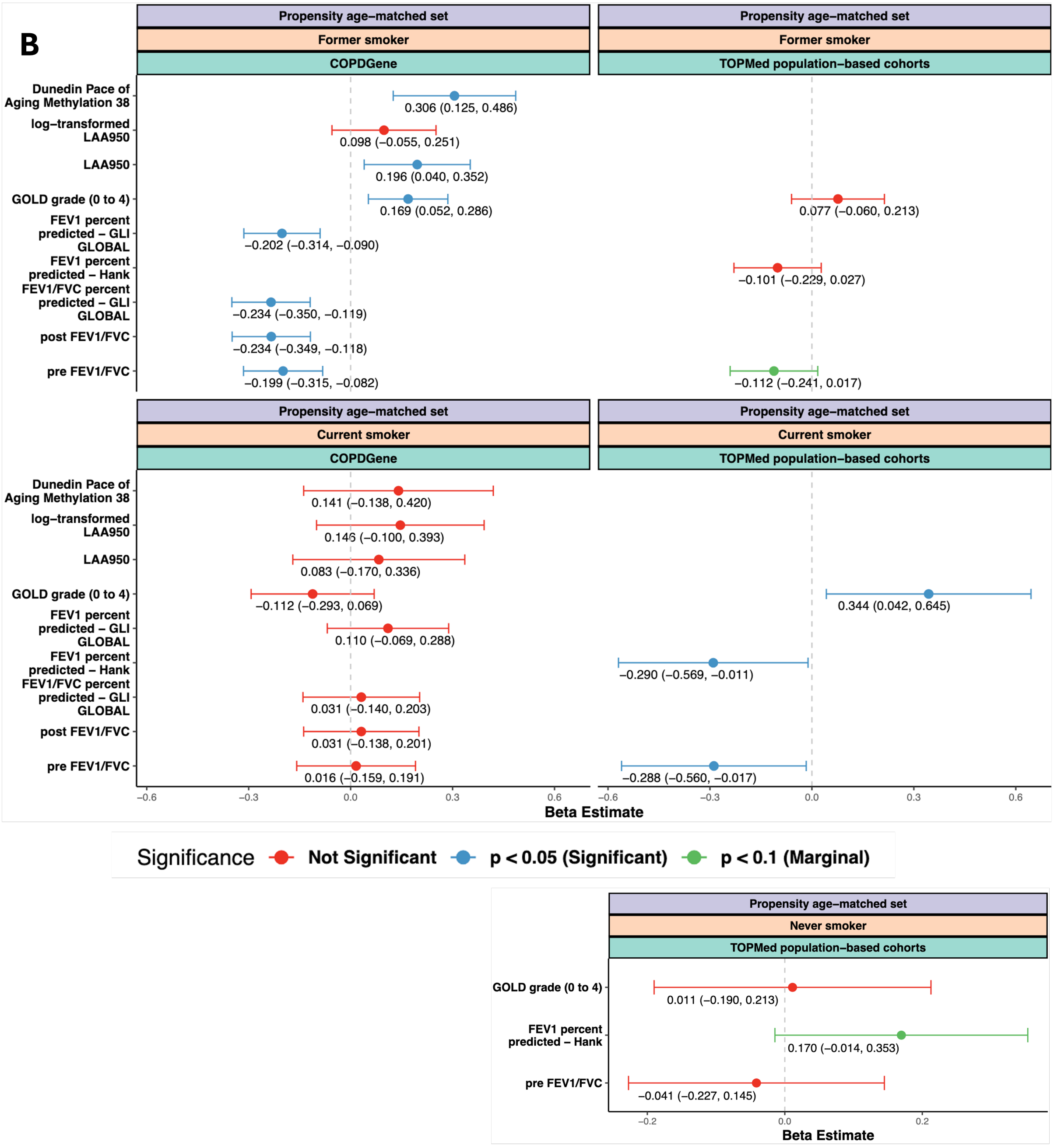


Supplementary Figure 5 Forest plots displaying A) odds ratios (OR) for categorical traits and B) beta coefficients for continuous traits with 95% confidence intervals (CI) in COPDGene and additional TOPMed population-based cohorts' smoking status stratification propensity aged-matched sets respectively. These are shown for both genetic ancestry principal components (PCs) adjustment, along with other confounders listed in the results section. Significance is color-coded.

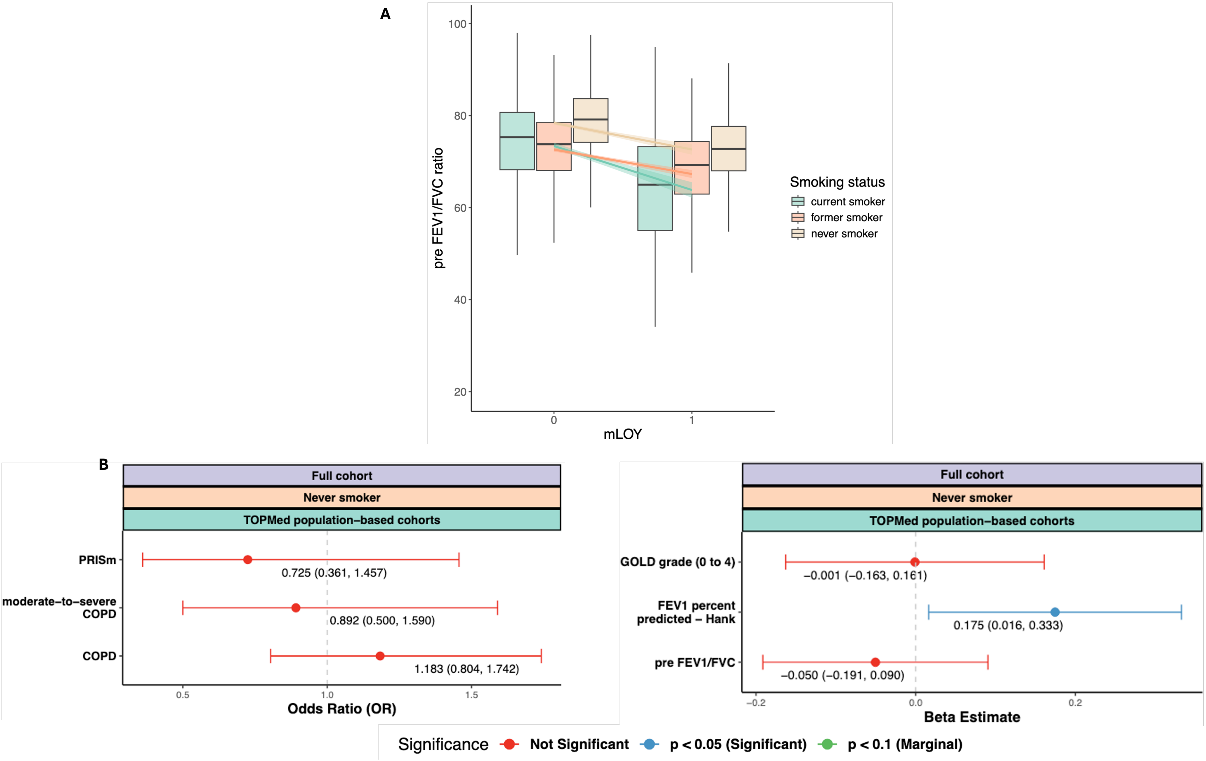


Supplementary Figure 6 A) Boxplot illustrating lung function (Y-axis) by mLOY status (X-axis), split by smoking status (interaction) in additional TOPMed population-based cohorts. The plot displays the distribution of lung function measures for both groups of individuals with mLOY and without mLOY, with separate boxes for current and former and never smoker, highlighting the combined effects of mLOY and smoking status on lung function. mLOY status is indicated as 0 for individuals without mLOY and 1 for individuals with mLOY. B) Forest plots illustrating beta coefficients with 95% confidence intervals (CIs), from the additional TOPMed population-based cohorts cross-sectional smoking status stratification analyses for never smoker group while both former and current smoker can be found in main Figure 3. The analyses are adjusted for genetic ancestry principal components (PCs) and additional confounders described in the results section. Statistically significant results are highlighted with color coding. Note that COPD, moderate COPD, severe COPD and PRISm are binary traits.

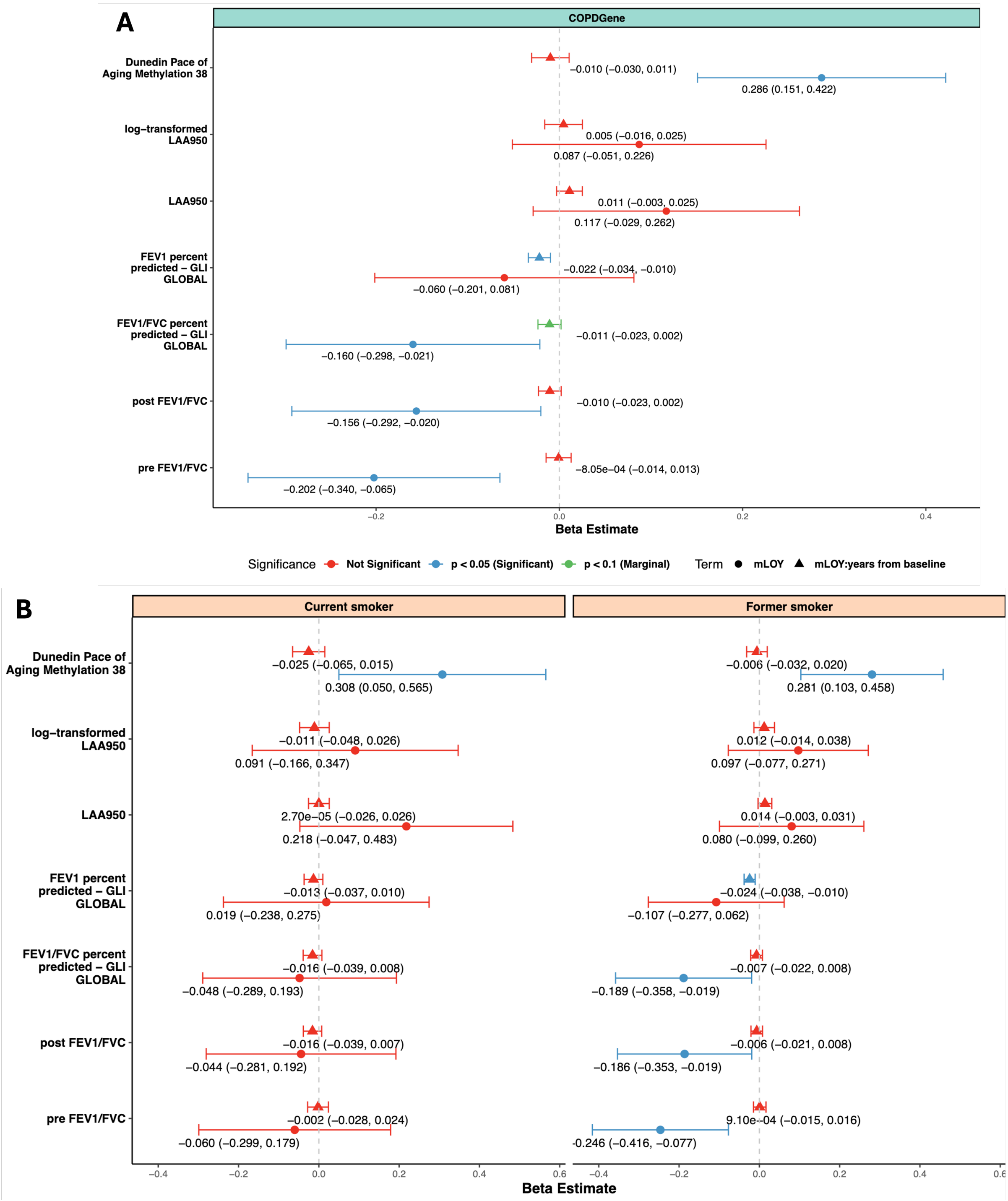


Supplementary Figure 7 Forest plots displaying beta coefficients for continuous traits with 95% confidence intervals (CI) in COPDGene longitudinal analysis. These are shown for both genetic ancestry principal components (PCs) adjustment, along with other confounders listed in the results section. Significance is color-coded.

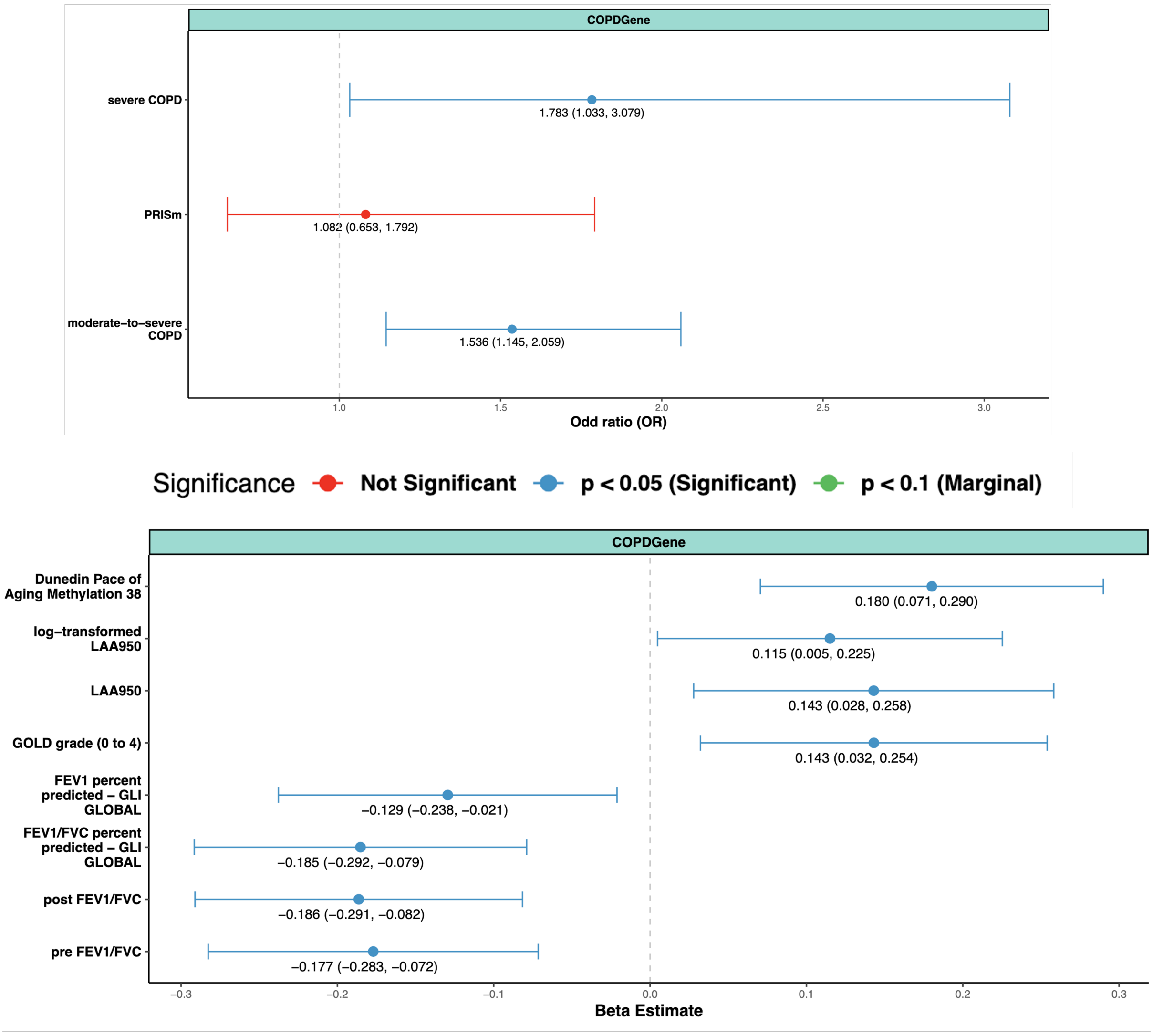


Supplementary Figure 8 Forest plots displaying odds ratios (OR) for categorical traits and beta coefficients for continuous traits, each with 95% confidence intervals (CI), in the COPDGene prospective analysis using full cohorts. The plots include adjustments for both genetic ancestry principal components (PCs), along with other confounders listed in the results section. Significance is color-coded. **Note:** The COPD outcome model failed to converge, due to multi-collinearity of the batch pc covariates. If we remove that the colinear batch PC 8, the model converges, and the effect size is as follows: effect size = 1.551, 95% CI 1.174 – 2.048, *p* = 0.002.

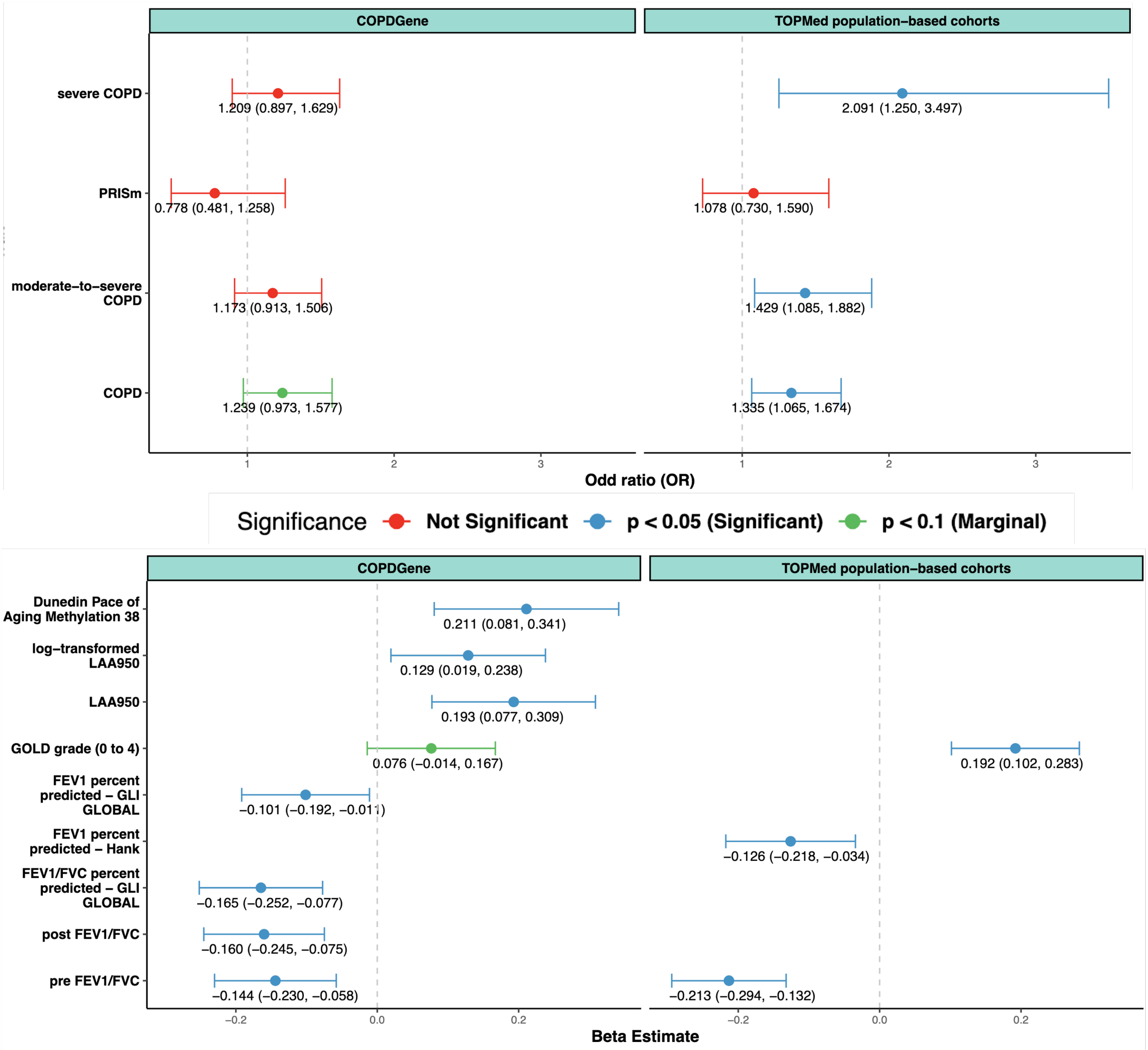


Supplementary Figure 9 Forest plots showing Odds ratios for categorical traits, and Beta coefficients with 95% CIs from COPDGene and additional TOPMed population-based cohorts respectively in cross-sectional sensitivity analysis with CF ≥ 10% for individuals with mLOY. Note that COPD, moderate COPD, severe COPD and PRISm are binary traits.

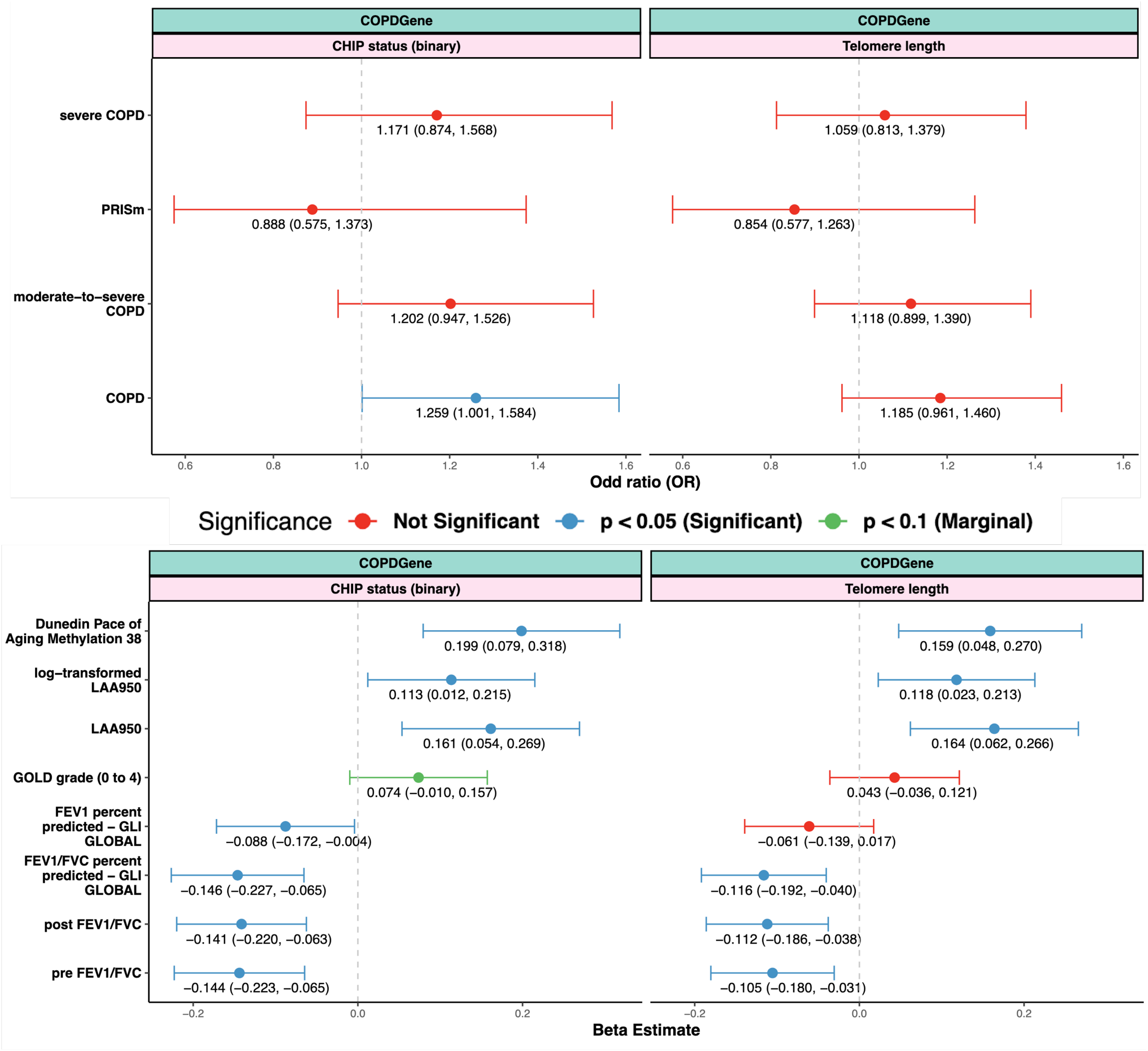


Supplementary Figure 10 Forest plots showing Odds ratios for categorical traits, and Beta coefficients with 95% CIs from COPDGene in cross-sectional sensitivity analysis by adding additional confounder CHIP status and Telomere length respectively. Note that COPD, moderate COPD, severe COPD and PRISm are binary traits.
